# Standardised Human Phenotype Ontology Annotation Enables High Quality Phenotypic Data Capture in a Real-World Common Variable Immunodeficiency Cohort

**DOI:** 10.64898/2026.04.29.26350963

**Authors:** Luiza C. Campos, Émeline Favreau, Daniel Greene, Justyna Blach, Moira Thomas, Khuloud Alsehaim, Leman Mutlu, Sara Elhadari, Archana Herwadkar, Julia Payne, Charley Lever, Dina Mahmoud, Fernando Moreira, Mary O’Sullivan, Maxine Berry, George Twigg, Alice C.J. Hart, Nehal Joshi, Stewart Fuller, INTREPID Consortium, Kenneth G.C. Smith, Ernest Turro, Matthew C. Cook, Chris Wallace, Siobhan O. Burns

**Affiliations:** University College London Institute of Immunity and Transplantation; Department of Immunology, Royal Free London NHS Foundation Trust; Cambridge Institute for Therapeutic Immunology and Infectious Diseases, Department of Medicine, University of Cambridge; Icahn School of Medicine at Mount Sinai, NY; Queen Elizabeth University Hospital Glasgow; University Hospitals Birmingham NHS Foundation Trust; University Hospitals of North Midlands; Salford Care Organisation, Northern Care Alliance NHS Trust; Leeds Teaching Hospitals NHS Trust; Imperial College Healthcare NHS Trust; The Walter and Eliza Hall Institute of Medical research (WEHI), Parkville, VIC, Australia; Department of Medical Biology, University of Melbourne, Melbourne, VIC, Australia; MRC Biostatistics Unit, University of Cambridge

**Keywords:** Common Variable immunodeficiency, CVID, Human phenotype Ontology, HPO, Phenotype, Primary Immunodeficiency, Biomarkers, data standardisation

## Abstract

**Background:** Patients with Common Variable Immunodeficiency (CVID) exhibit diverse clinical manifestations, indicating heterogeneity in pathogenic mechanisms. Systematic application of standardised phenotyping in large cohorts is essential to dissect this heterogeneity. The Human Phenotype Ontology (HPO) provides a structured framework for capturing and comparing disease phenotypes.

**Objective:** To evaluate the implementation and outcomes of HPO-based phenotyping in CVID patients enrolled for whole-genome sequencing in a large national adult primary immunodeficiency cohort.

**Methods:** We developed a web-based Phenotype Capture Tool and delivered structured clinician training to standardise HPO annotation. Numerical laboratory parameters were mapped to corresponding HPO terms to enrich patient records.

**Results:** We coded the phenotypes of 526 CVID patients across 11 UK centres. Clinician training increased phenotype granularity and improved phenotyping consistency between clinicians. We assigned 883 unique HPO terms across the cohort and applied logical rules to the terms to classify patients into an infection-only group and a complex phenotype group (42% vs 58%, respectively). Patients in the complex phenotype group were significantly more likely to have reduced switched memory and expanded CD21^low^ B cells, as well as pathogenic variants in IUIS-listed genes overall and pathogenic *NFKB1* variants specifically. Having a pathogenic variant in an IUIS-listed gene was associated with *Autoimmune hemolytic anemia* and having a pathogenic *NFKB1* variant specifically was associated with *Autoimmune neutropenia*.

**Conclusion:** This is the first study to systematically collect granular HPO-coded phenotypes in a large real-world CVID cohort, refining the CVID landscape and providing a comprehensive CVID HPO term set relevant for international research.

**Clinical Implication:** HPO allows systematic capture of CVID phenotypes with low inter-clinician variability and improves comparison of cohorts, enhancing identification of disease heterogeneity essential to support genotype-phenotype studies and targeted therapeutic strategies.

**Capsule summary:** HPO-based phenotyping of 526 CVID patients improved annotation quality, identifying immunological and genetic associations with clinical manifestations, distinguishing infection-only from complex disease and refining clinical characterisation to support international collaboration.

## Introduction

Common variable immunodeficiency (CVID) is the most prevalent primary immunodeficiency (PID), requiring lifelong care. Affected individuals display heightened susceptibility to infection, and some patients additionally experience autoimmunity, inflammation, lymphoproliferation and malignancy ^1–4^. Delineating these subgroups is essential, as they carry different prognostic and therapeutic implications ^5^. Although many disease-associated genes have now been identified in CVID, overall, monogenic causes account for only 25-30% of cases, underscoring the mechanistic heterogeneity and etiological knowledge gaps of CVID^6–8^.

Existing CVID classification systems are based on B cell phenotyping ^9–11^. These approaches have identified associations between B cell abnormalities and specific clinical patterns, but rely on clinical information that is often incomplete or inconsistent. A deeper and standardised phenotypic description has the potential to refine patient stratification and uncover the biological basis of CVID heterogeneity.

Among available standardised medical terminologies, the Human Phenotype Ontology (HPO) offers the most detailed and structured framework for describing human disease phenotypes. It captures hierarchical relationships between over 18,000 phenotypic features at multiple level of abstraction. HPO is particularly well-suited to rare diseases as it supports powerful computational applications, including semantic similarity analyses, clustering, and machine learning for diagnosis and variant prioritisation ^12–20^. Importantly, HPO accommodates large, heterogeneous datasets, making it valuable for resolving the phenotypic complexity of disorders such as CVID.

The effectiveness of HPO-based approaches depends heavily on the quality and completeness of the annotations provided. Barriers at the clinical service level – limited familiarity with ontologies, time pressure, and variable coding practices – reduce data fidelity. Recent initiatives, including incorporation of HPO terminology into inborn errors of immunity (IEI) working definitions within the European Society for Immunodeficiencies registry (ESID-R), and coordinated efforts by the European Reference Network on Rare Primary Immunodeficiency, Autoinflammatory and Autoimmunity Diseases (ERN-RITA) with ESID to expand and reannotate IEI-relevant terms, have begun to address these gaps ^21–23^. However, for many IEIs, the HPO remains under curated or applied inconsistently, leading to sparse or inaccurate capture of disease-specific manifestations and their frequencies ^22^. To date, no studies have evaluated the feasibility and yield of HPO-phenotyping of large real-world IEI cohorts.

To evaluate whether a systematic, clinician-driven workflow can achieve high-quality HPO annotation in CVID, and if so, whether such annotations can uncover clinically useful patterns, we leveraged the INTREPID (Integrated Translational Research in Primary Immunodeficiency) project, a UK-wide whole-genome sequencing (WGS) initiative for IEI patients ^24^. We developed a web-based Phenotype Capture Tool (PCT) and delivered clinician training to maximise the specificity and completeness of HPO coding. By improving clinicians’ familiarity with HPO and providing practical tools for data entry, we aimed to increase annotation accuracy that would ultimately allow more granular dissection of CVID heterogeneity to support patient stratification and informed therapeutic decision-making.

## Methods

### Patient recruitment and Data Collection

We collected data under the NIHR BioResource Rare Diseases ethics infrastructure and clinicians recruited patients to the NIHR BioResource (https://bioresource.nihr.ac.uk/studies/nbr120/). Patients above the age of 16 years, with suspected primary immunodeficiency and/or recurrent or unusual infections were eligible for the study. Patients with secondary immunodeficiencies were excluded.

Although recruitment is ongoing, analyses were performed on a database freeze in September 2025. Records with a top-level diagnosis of CVID assigned by their treating clinician according to ESID criteria^23^ were included.

To support systematic phenotypic data collection in this cohort, we developed the Phenotype Capture Tool (PCT) within the INTREPID project to replace paper-based data collection and facilitate entry of HPO terms and laboratory values for participants. See Online Repository Methods for details.

### Clinician training

Clinicians at each recruiting centre were trained to collect HPO terms describing IEI patients, and how to use the PCT. All clinicians practiced on the same clinical case scenarios, with data stored in a training instance of the PCT. See Online Repository Methods for details.

### Training data analysis

We assessed the impact of clinician training by measuring pairwise Lin’s semantic similarity scores (0 = not similar, 1 = highly similar) using the ontologyIndex v2.5 R package ^25^ between sets of HPO terms assigned to a given test case annotated by ten different clinicians in the training instance of the PCT. Higher similarity scores for a given case were interpreted as greater consistency of phenotype coding between clinicians.

To evaluate changes in annotation completeness, we compared the number of nonredundant HPO terms recorded for each participant at the time of their initial enrolment for WGS and after completion of the PCT training. Annotation granularity was assessed by comparing the median and interquartile range (IQR) of nonredundant HPO terms per patient before and after training, and differences in distributions were tested using the Kolmogorov-Smirnov test (*P* < 0.05 considered significant). Similarity analysis between pre-and post-training HPO annotations for paired entries was done by calculating Lin’s similarity. This analysis is based on the information content for the HPO terms relative to their common ancestor.

### Patient categorisation

Phenotypic abnormalities were categorised based on the presence of HPO terms, after propagation of terms implied by the HPO’s *is-a* graph structure, based on a phenotype-defining set of HPO terms (see Online Repository Methods). Presence or absence of granulomatous lymphocytic interstitial lung disease (GLILD) was recorded through a yes/no box in the PCT as GLILD is not an HPO term.

Patients were subsequently classified into broad clinical groups-– infection only or complex group – based on the presence or absence of non-infectious complications.

In addition to clinical annotations, laboratory measurements such as B cell counts, immunoglobulin levels, and other results were systematically mapped to corresponding HPO terms to enrich the dataset (See Online Repository Methods and Table E1 in the Online Repository).

Frequencies of phenotypic abnormalities were compared across patient groups stratified by B cell subset counts (according to the EUROClass system ^11^), immunoglobulin levels, and diagnostic genetic variants^26^.

Patient characteristics (e.g. B cell counts) were not uniformly distributed across centres. We accounted for between-centre variation using the Cochran-Mantel-Haenszel test. Associations were tested between patient grouping based on their B cell counts, immunoglobulin levels, genetic variants and phenotypes (key HPO groups, individual HPO terms, and presence of GLILD. We corrected for multiple testing using the Benjamini-Hochberg false discovery rate (BH-FDR) method, considering BH adjusted *P* < 0.05 significant. All analyses were performed in R (version 4.3.2).

### Statistical analysis

In addition to the statistical analyses described in the sections above, the analysis pipeline is available in this GitHub repository (https://github.com/EmelineFavreau/Analysing-CVID-HPO).

## Results

### Clinician Training significantly improved the granularity and consistency of phenotype annotation

At the time of the data freeze, 28 clinicians across 16 centres had completed training. From 1,406 records compiled from these centres for patients with PID (see Table E2 in the Online Repository for full diagnostic distribution), 526 CVID records annotated after clinician training were included in the analyses. These records were contributed by 11 centres (see Online Repository Methods).

To evaluate the effect of HPO training, 10 clinicians annotated the same clinical vignette using a mock version of the PCT. Pairwise comparisons of resulting HPO term sets showed high consistency across clinicians, with most producing highly similar annotations (median, 0.80; IQR, 0.17) (Figure 1a). Most clinician pairs achieved semantic similarity scores above 0.5, indicating concordant coding practices. The main deviations were observed for clinican#2, and to a lesser extent clinician#9, whose annotations showed lower similarity relative to the rest of the group, reflecting individual coding choices rather than systematic variability.

**Figure 1:**
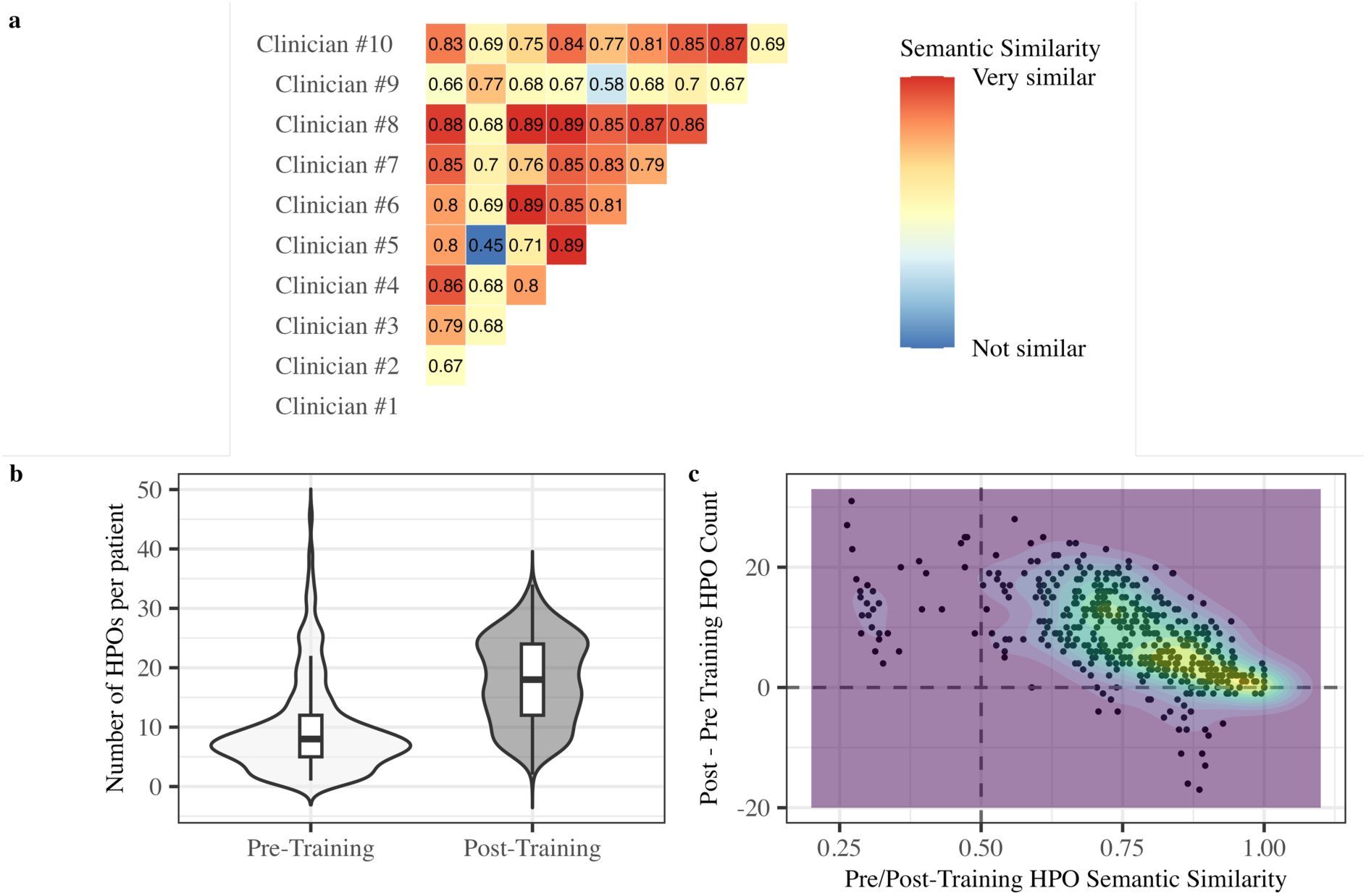
Clinician training enhances consistency and granularity of HPO annotations. a) Pairwise semantic similarity (0-1 scale) between HPO term set used by clinicians, independently annotating the same clinical case following HPO training. Each row/column represents a clinician. All but one comparison (44/45) indicate high concordance (≥0.5); b) Number of HPO terms per patient before and after clinician training. Training significantly increased the number of captured phenotypic terms per patient from a median of 7 to 19; IQR 7 to 12; P < 2.2×10⁻¹⁶, Kolmogorov-Smirnov test; c) Change in number and similarity of HPO terms per patient before and after clinician training. Each point corresponds to one patient.The density of the points is colour-coded, with warmer colour demonstrating higher density. Training increased the number of HPO terms annotations per patient.

We assessed the effect of training on real patient annotations by comparing two annotation sets: 1) pre-training annotations, entered at WGS recruitment, before clinicians received access to the PCT with formal HPO training; and 2) post-training annotations, entered when clinicians subsequently reviewed the same records after completing HPO/PCT training. Among 487 cases with paired entries, we observed a significant increase in the number of nonredundant HPO terms annotated per patient, from a median of 7 (IQR = 8) before training to 19 (IQR = 11) after training, with the distributions differing significantly (Kolmogorov-Smirnov *P* < 2.2 x 10⁻¹⁶; Figure 1b).

Direct comparison of pre-and post-training HPO annotations demonstrated an increase in phenotypic granularity, with most patients receiving additional non-redundant terms after training. In a small subset (n = 34), the total number of post-training terms recorded was lower than pre-training, reflecting refinement and replacement of high-level or non-specific terms with fewer, more informative descriptors rather than loss of phenotypic information. Overall, semantic similarity between pre-and post-training annotation sets was high (mean similarity 0.77; IQR 0.19). When examined by similarity sections, 93% of records showed substantial similarity (> 0.5), including 46% that were highly similar (> 0.8), indicating preservation of each patient’s core phenotypic profile. Consistent with this interpretation, post-training annotations frequently retained core concepts while adding specificity, for example maintaining *Autoimmunity* [HP:0002960] alongside addition of *Autoimmune thrombocytopenia* [HP:0001973] and *Autoimmune haemolytic anaemia* [HP:0001890] and incorporating laboratory-specific features such as *Increased transitional B cells proportion* [HP:0030381]. Only 7% of records had lower similarity values (< 0.5). In these cases, pre-training annotations consisted of very few HPO terms (1-4 terms), whereas post-training annotations were considerably more extensive (8-30 terms) (Figure 1c).

### Cohort Overview

Among the 526 CVID records, 275 were female and 243 were male patients (sex was not recorded for 8 individuals), and a median age of 58 years (Table E3 in the Online Repository).

Immunoglobulin levels were available for 500 patients (95%). The majority, (n = 410, 78%) had panhypogammaglobulinemia while 65 patients (12.4%) had low IgG and IgA with normal IgM, and 13 patients (2.5%) had low IgG and IgM with normal IgA. Few patients (n = 12, 2.3%) had low IgG and IgA with elevated IgM (Table I).

**Table I:** Cohort overview. The table summarises the frequency of patients with each clinical phenotype based on HPO term sets and displays the distribution of immunoglobulin level patterns in the INTREPID cohort (526 CVID patients).

| **Clinical Phenotype** | **Number of Records, %** |
| --- | --- |
| Splenomegaly | 145, 27.6% |
| Lymphadenopathy | 99, 18.8% |
| Granulomatous disease | 53, 10.1% |
| Autoimmunity | 153, 29.1% |
| .... of which are Autoimmune cytopaenias | 105, 68.6% (out of 153) |
| NRH liver | 37, 7% |
| Gastrointestinal inflammation | 52, 9.9% |
| Hematological neoplasm | 40, 7.6% |
| **Immunoglobulin subgroups** |  |
| lowIgG_lowIgA_lowIgM | 410, 77.95% |
| lowIgG_lowIgA_normalIgM | 65, 12.36% |
| lowIgG_normalIgA_lowIgM | 13, 2.47% |
| lowIgG_lowIgA_highIgM | 12, 2.28% |

Switched memory B cell measurements were available for 363 (69%) participants, with 183 (50.4%) classified as smB+ and 180 (49.6%) as smB-. CD21^low^ B cells were recorded for 293 (55.7%), of whom 148 (50.5%) had expansion of this subset and 145 (49.5%) had normal CD21^low^ levels. Transitional B cells as proportion of B cells were recorded for 289 patients (54.9%), with 263 (91%) within the normal range and 26 (9.0%) with elevated values.

Categorisation according to EUROClass definitions identified 180 patients in the smB-group, with further subdivision into smB-/CD21^lo^ (n = 75), smB-/CD21^norm^ (n = 67), smB-/Tr^hi^ (n = 10), and smB-/Tr^norm^ (n = 129). Among 183 patients with smB+, 70 were classified as smB+/CD21^lo^ and 78 as smB+/CD21^norm^. Compared with the EUROClass cohort ^11^, the INTREPID cohort showed similar proportions of patients classified as smB+CD21^norm^ and smB+CD21^lo^ (Chi-square test *P* = 0.032) and a smaller proportion of patients classified as smB-Tr^hi^ (Chi-square test *P* = 0.007) (Figure 2a and b).

**Figure 2:**
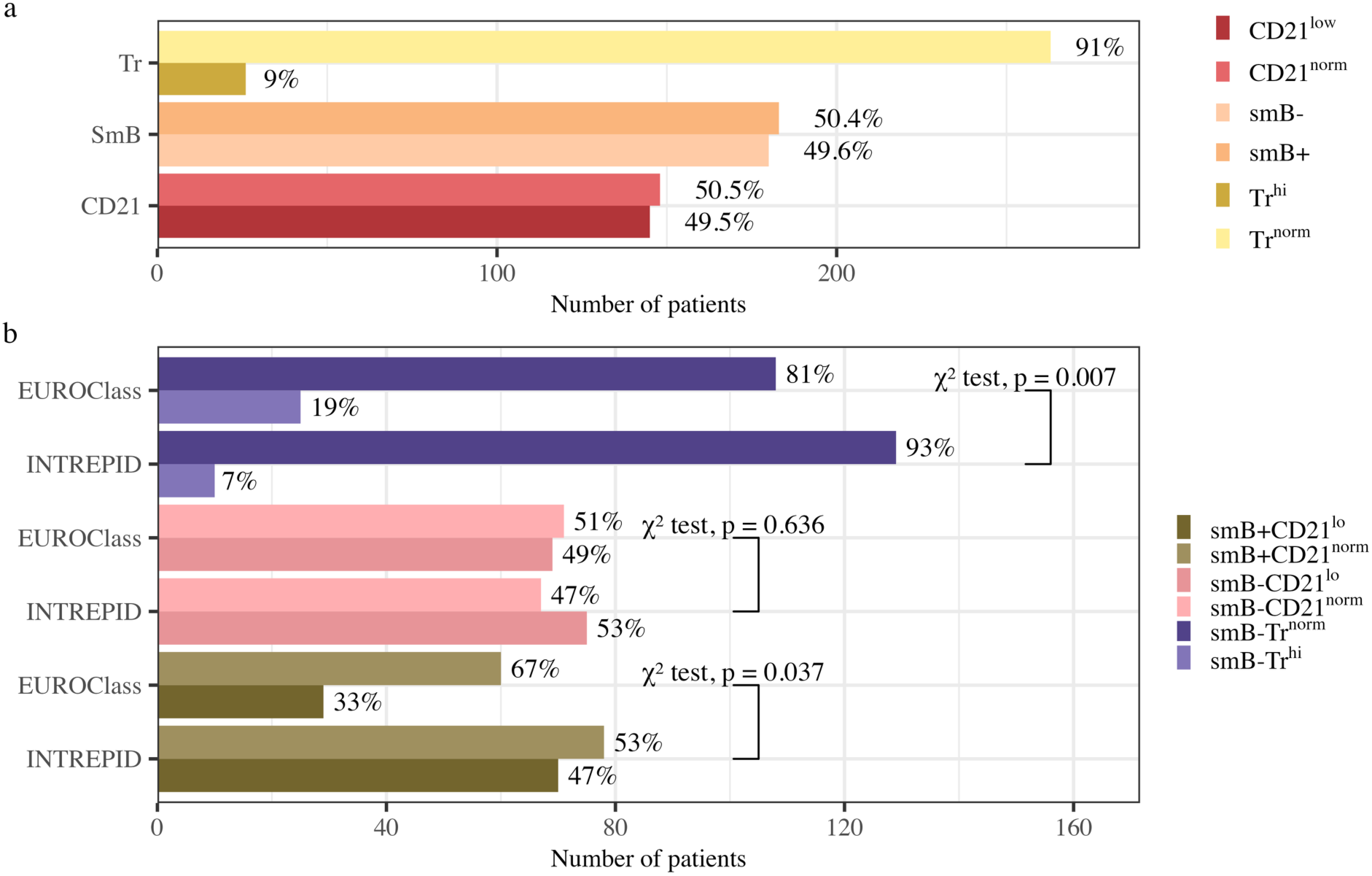
Significant differences in smB/CD21 and smB/Tr frequencies between INTREPID and EUROClass cohorts. a) Number of INTREPID patients for single markers: CD21^norm^ (CD21^low^ <10 %) or CD21^low^ (patients with expanded CD21^low^ subset ≥ 10%) out of 293 with CD21 measured; smB-(≤ 2% smB) or smB+ (> 2%); transitional B cell Tr^norm^ (<9%) or Tr^hi^ (≥ 9%) out of 363 with smB measured; b) Distribution of B cell subsets defined by EUROClass system in both INTREPID and EUROClass cohorts. INTREPID shows similar proportions of smB+CD21^norm^ vs smB+CD21 ^lo^ and smaller proportion of patients with smB-Tr^hi^ compared with smB-Tr^norm^.

WGS results were available for 433 patients (82.3%), of whom 94 (17.9%) carried an IUIS genetic variant. Among these, 19 patients (20.2% of diagnosed, 4.4% of sequenced) carried *NFKB1* variants. Variants in *TNFRSF13B* (encoding TACI) were identified in 37 patients (8.5% of sequenced), including 27 (6.2%) with a so-called canonical variant (either p.Arg181Glu or p.Cys104Arg), and 9 (2.1%) with a rare variant. The remaining 40 patients (42.6% of those with a confirmed diagnosis) carried variants in genes other than *NFKB1* or *TNFRSF13B* (Table II).

**Table II:** Genetic variants identified in the INTREPID CVID cohort. There are 80 patients with a genetic diagnosis.

| Patient | Gene | cDNA | Protein | Sex |
| --- | --- | --- | --- | --- |
| 1 | BCL11B | c.1735dup | p.(Ala579GlyfsTer11) | M |
| 2 | BTK | c.837T>A | p.(Tyr279Ter) | M |
| 3 | CD40LG | c.98T>A | p.(Ile33Asn) | M |
| 4 | CTLA4 | chr2-203867482-203869297 | Unknown | M |
| 5 | CTLA4 | c.209G>A | p.(Arg70Gln) | M |
| 6 | CXCR4 | c.1003G>A | p.(Gly335Ser) | M |
| 7 | GATA2 | c.599G>C | p.Gly200Ala | M |
| 8 | ICOS | c.58+1G>A | Unknown | M |
| 9 | IKZF1 | c.500A>G | p.(His167Arg) | F |
| 10 | LRBA | c.8422G>T | p.(Gly2808Ter) | M |
| 11 | MRE11 | c.1516G>T | p.(Glu506Ter) | M |
| 12 | NBAS | c.7041_7043del | p.(Leu2348del) | M |
| 13 | NFKB1 | c.187delG | p.(Glu63LysfsTer64) | F |
| 14 | NFKB1 | DEL 4:103370996-103528207 | Unknown | F |
| 15 | NFKB1 | c.1005delG | p.(Arg336GlyfsTer96) | F |
| 16 | NFKB1 | c.1621_1622delGA | p.(Asp541Ter) | F |
| 17 | NFKB1 | c.293T>A | p.(Val98Asp) | F |
| 18 | NFKB1 | c.295C>T | p.(Gln99Ter) | M |
| 19 | NFKB1 | c.260T>G | p.(Ile87Ser) | F |
| 20 | NFKB1 | Del 4:102515817-102731498 | Unknown | F |
| 21 | NFKB1 | Unknown | Unknown | F |
| 22 | NFKB1 | c.904dupT | p.(Ser302PhefsTer7) | F |
| 23 | NFKB1 | c.2335dup | p.(Met779AsnfsTer9) | M |
| 24 | NFKB1 | c.1423delG | p.(Ala475ProfsTer10) | M |
| 25 | NFKB1 | c.160-1G>A | p.(Arg54_Lys86del) | F |
| 26 | NFKB1 | DUP 4:103433198-103487573 | Unknown | M |
| 27 | NFKB1 | c.1901dup | p.(Leu636ThrfsTer11) | F |
| 28 | NFKB1 | c.2378C>G | p.(Pro793Arg) | F |
| 29 | NFKB1 | c.830dupA | p.(Lys278GlufsTer3) | F |
| 30 | NFKB1 | c.1968_1971dup | p.(Leu658SerfsTer8) | F |
| 80 | NFKB1 | Unknown | Unknown | F |
| 31 | NFKB2 | c.1469+1G>T | Unknown | M |
| 32 | NFKB2 | c.1903C>T | p.(Arg635Ter) | F |
| 33 | NHEJ1 | c.170G>A | (p.Arg57Gln) | M |
| 34 | PIK3CD | c.427A>G | p.(Met143Val) | M |
| 35 | PIK3R1 | c.1422_1425+1del | Unknown | M |
| 36 | PIK3R1 | c.1425+1G>A | Unknown | F |
| 37 | RAC2 | c.106G>A | p.(Val36Met) | M |
| 38 | RAG1 | c.334C>T | p.(Arg112Cys) | F |
| 39 | SOCS1 | c.476_480dup | p.(Met161AlafsTer46) | F |
| 40 | STAT1 | c.1154C>T | p.(Thr385Met) | F |
| 41 | STAT3 | c.812C>T | p.(Ala271Val) | F |
| 42 | STAT3 | c.1145G>A | p.(Arg382Gln) | F |
| 43 | TNFRSF13B | c.542C>A | p.(Ala181Glu) | F |
| 44 | TNFRSF13B | c.542C>A | p.(Ala181Glu) | M |
| 45 | TNFRSF13B | c.542C>A | p.(Ala181Glu) | F |
| 46 | TNFRSF13B | c.542C>A | p.(Ala181Glu) | F |
| 47 | TNFRSF13B | c.542C>A | p.(Ala181Glu) | F |
| 48 | TNFRSF13B | c.542C>A | p.(Ala181Glu) | M |
| 49 | TNFRSF13B | c.542C>A | p.(Ala181Glu) | F |
| 50 | TNFRSF13B | c.542C>A | p.(Ala181Glu) | M |
| 51 | TNFRSF13B | c.310T>C | p.(Cys104Arg) | F |
| 52 | TNFRSF13B | c.310T>C | p.(Cys104Arg) | F |
| 53 | TNFRSF13B | c.310T>C | p.(Cys104Arg) | M |
| 54 | TNFRSF13B | c.310T>C | p.(Cys104Arg) | M |
| 55 | TNFRSF13B | c.310T>C | p.(Cys104Arg) | F |
| 56 | TNFRSF13B | c.310T>C | p.(Cys104Arg) | F |
| 57 | TNFRSF13B | c.310T>C | p.(Cys104Arg) | M |
| 58 | TNFRSF13B | c.310T>C | p.(Cys104Arg) | F |
| 59 | TNFRSF13B | c.310T>C | p.(Cys104Arg) | M |
| 60 | TNFRSF13B | c.310T>C | p.(Cys104Arg) | F |
| 61 | TNFRSF13B | c.310T>C | p.(Cys104Arg) | F |
| 62 | TNFRSF13B | c.310T>C | p.(Cys104Arg) | F |
| 63 | TNFRSF13B | c.310T>C | p.(Cys104Arg) | F |
| 64 | TNFRSF13B | c.310T>C | p.(Cys104Arg) | F |
| 65 | TNFRSF13B | c.310T>C | p.(Cys104Arg) | F |
| 66 | TNFRSF13B | c.310T>C | p.(Cys104Arg) | F |
| 67 | TNFRSF13B | c.310T>C | p.(Cys104Arg) | F |
| 68 | TNFRSF13B | c.310T>C | p.(Cys104Arg) | M |
| 69 | TNFRSF13B | c.310T>C | p.(Cys104Arg) | F |
| 70 | TNFRSF13B | c.310T>C | p.(Cys104Arg) | F |
| 71 | TNFRSF13B | c.310T>C | p.(Cys104Arg) | M |
| 72 | TNFRSF13B | c.310T>C | p.(Cys104Arg) | F |
| 73 | TNFRSF13B | c.733G>A | p.(Ala245Thr) | M |
| 74 | TNFRSF13B | c.365G>A | p.(Arg122Gln) | M |
| 75 | TNFRSF13B | c.605G>A | p.(Arg202His) | F |
| 76 | TNFRSF13B | c.515G>A | p.(Cys172Tyr) | M |
| 77 | TNFRSF13B | c.832G>A | p.(Gly278Ser) | F |
| 78 | TNFRSF13B | c.227_231del | p.(Gly76ValfsTer3) | F |
| 79 | TNFRSF13B | c.452C>T | p.(Pro151Leu) | F |

### HPO terms in CVID

Based on HPO-coded features, patients were classified into two broad phenotypic groups, according to a phenotype-defining set of HPO terms (see Online repository Methods): the infection-only group, consisting of patients with recurrent or chronic infections, with or without bronchiectasis and the complex group, consisted of patients with infections plus features of immune dysregulation, such as autoimmunity, splenomegaly, lymphoproliferation, granulomas, GLILD and/or nodular regenerative hyperplasia (NRH) of liver.

Using this system, 220 patients (41.8%) were assigned to the infection-only group, while 306 patients (58.2% were classified as having complex CVID.

To assess whether clinical subgroups differed in their total phenotypic profiles, we created HPO embeddings using node2vec and performed a principal component analysis (PCA). The first principal component captures differences between complex and infection-only patients, with the centre of mass of complex patients lying to the right of infection-only patients (Figure 3a). Variation between centres is also seen with centres contributing larger numbers of patients tending to fall to the right (Figure 3b).

**Figure 3:**
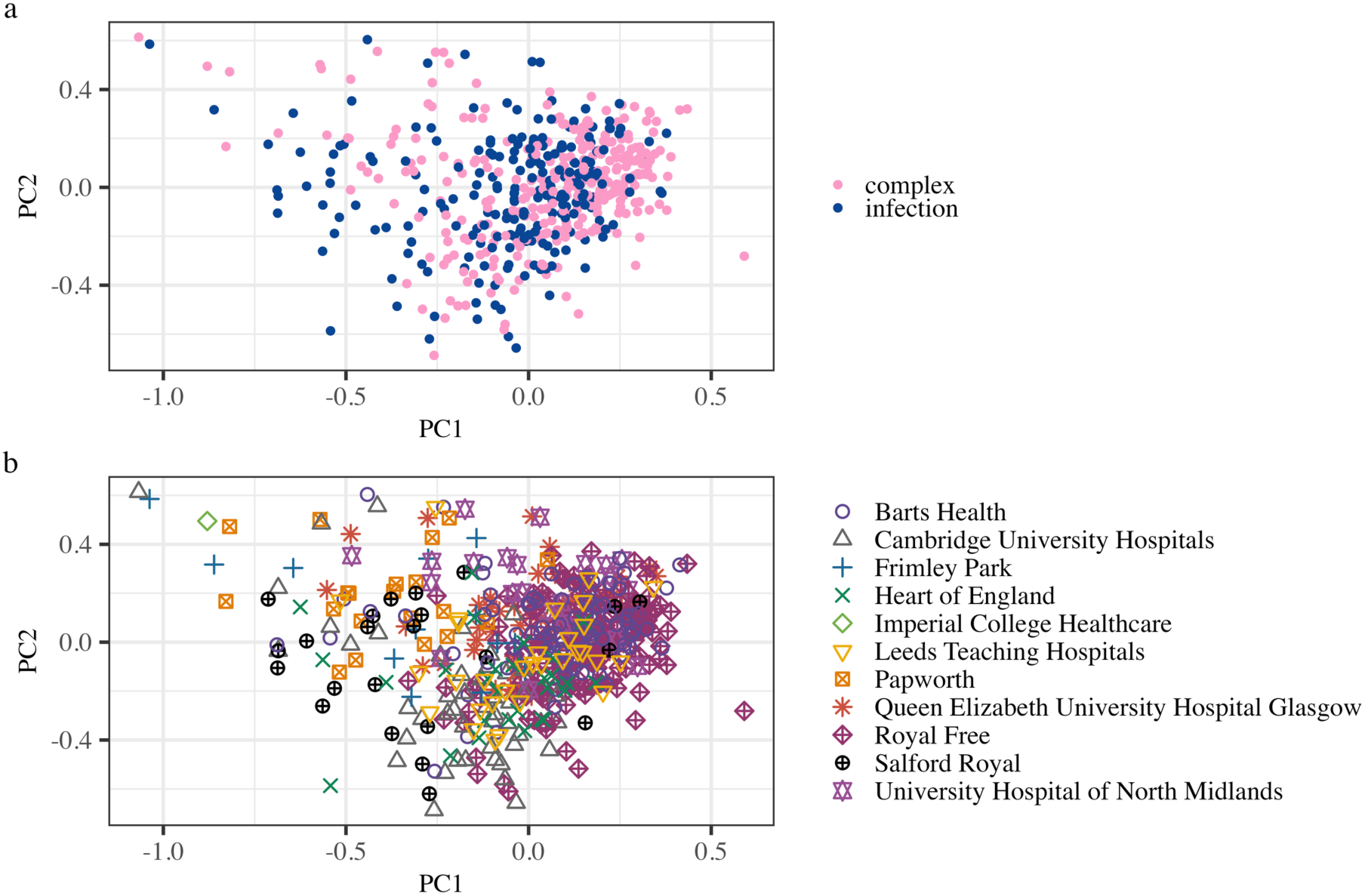
PCA of HPO terms captures both differences between complex and infection patients, and between patients from different centres. Each point represents an individual patient with their HPO term sets encoded using ontology-based embeddings. a) PCA coloured by clinical clustering: PC1 highlights differences between Complex CVID patients (right) and Infection/bronchiectasis patients (left). b) PCA coloured by referring center. Centers contribute larger numbers of patients skewing toward the right side of PC1.

A total of 883 unique HPO terms were used to describe the patients in our CVID cohort. Table E2 in the Online Repository lists all terms and their frequencies across the cohort.

As expected, features that form part of the diagnostic criteria for CVID were among the most frequently annotated phenotypes, including decreased circulating IgG, IgA and IgM levels (present in 80%-98% of the patients), decreased proportion of class-switched memory B cells (65%), and recurrent bacterial infections (38.4%). Importantly, beyond these core diagnostic features, we also captured a wide range of granular phenotypes, reflecting disease complexity, such as decreased DLCO (10%), ground-glass opacification (9.5%), nodular regenerative hyperplasia of liver (7%), portal hypertension (6.6%), and villous atrophy (3.2%).

When stratified into infection-only and complex groups, analysis of term frequencies (Figure 4 a and b) identified splenomegaly as the most frequent immune dysregulation HPO term in the complex group (44.8%). Autoimmune cytopenias, particularly *Autoimmune thrombocytopenia* [HP:0001973], were the most frequent autoimmunity-related terms observed in 49.5% and 27.5% of the patients in the complex group, respectively. No differences were observed in the infection-related specific terms when comparing between these groups. A small number of patients do not have *Decreased circulating IgG level* [HP:0004315] coded - where clinicians entered *Hypogammaglobulinemia* [HP:0004313] instead of coding each immunoglobulin subclass separately.

**Figure 4.**
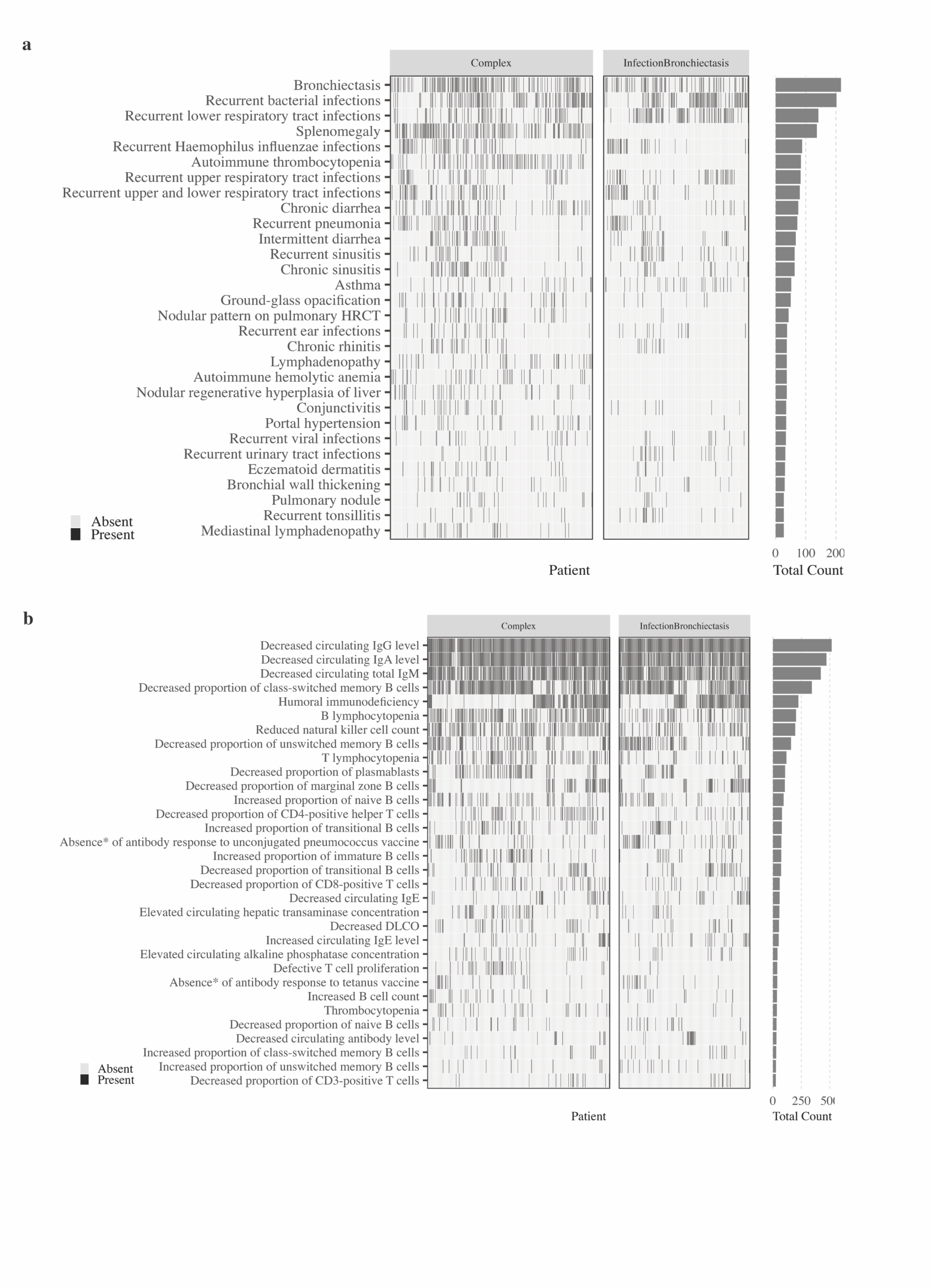
Presence and frequency of common clinical and laboratory HPO terms in 526 patients classified as Infection with/without bronchiectasis or complex CVID. Presence/absence heatmap of the 63 most frequent (>25 occurrences) clinical (panel a) and laboratory HPO terms (panel b), with terms in rows and individual patients in columns. Bar plots on the right of each panel show the frequency of each HPO term in the cohort. Note: * two long HPO terms are abbreviated: Complete or near-complete absence of specific antibody response to unconjugated pneumococcus vaccine [HP:0410300]; Complete or near-complete absence of specific antibody response to tetanus vaccine [HP:0410295].

### Identification of missing HPO terms

During curation, we identified phenotypic features absent from the current HPO. Most of these missing terms were related to the presence of granulomas in different organs, recurrent or chronic infections, including those caused by specific pathogens, and laboratory-based features, critical for characterising CVID patients. In total, 41 disease-specific HPO terms were identified. These terms have been entered into the formal HPO new-term submission pipeline. We also suggest additional term relationships, including adding new parent or children terms to existing terms and assigning terms frequencies (granuloma-related terms shown in Figure E2; all new terms and relationships suggestions on github).

### Immunological and Genetic Correlates of Disease Complexity

Next, we evaluated the utility of HPO-based data capture of clinical and laboratory features to identify specific disease associations. Complex disease was significantly associated with very low levels of switched memory B cells (<2%; BH adjusted *P* = 9×10^-5^) and with expansion of the CD21^low^ B cell subset (>10%; BH adjusted *P* = 5×10^-5^). In addition, *NFKB1* variants were only present among patients with complex disease (*P* = 0.0035), and the overall identification of diagnostic variants was also significantly enriched in this group (*P* = 0.002) (Figure 5).

**Figure 5:**
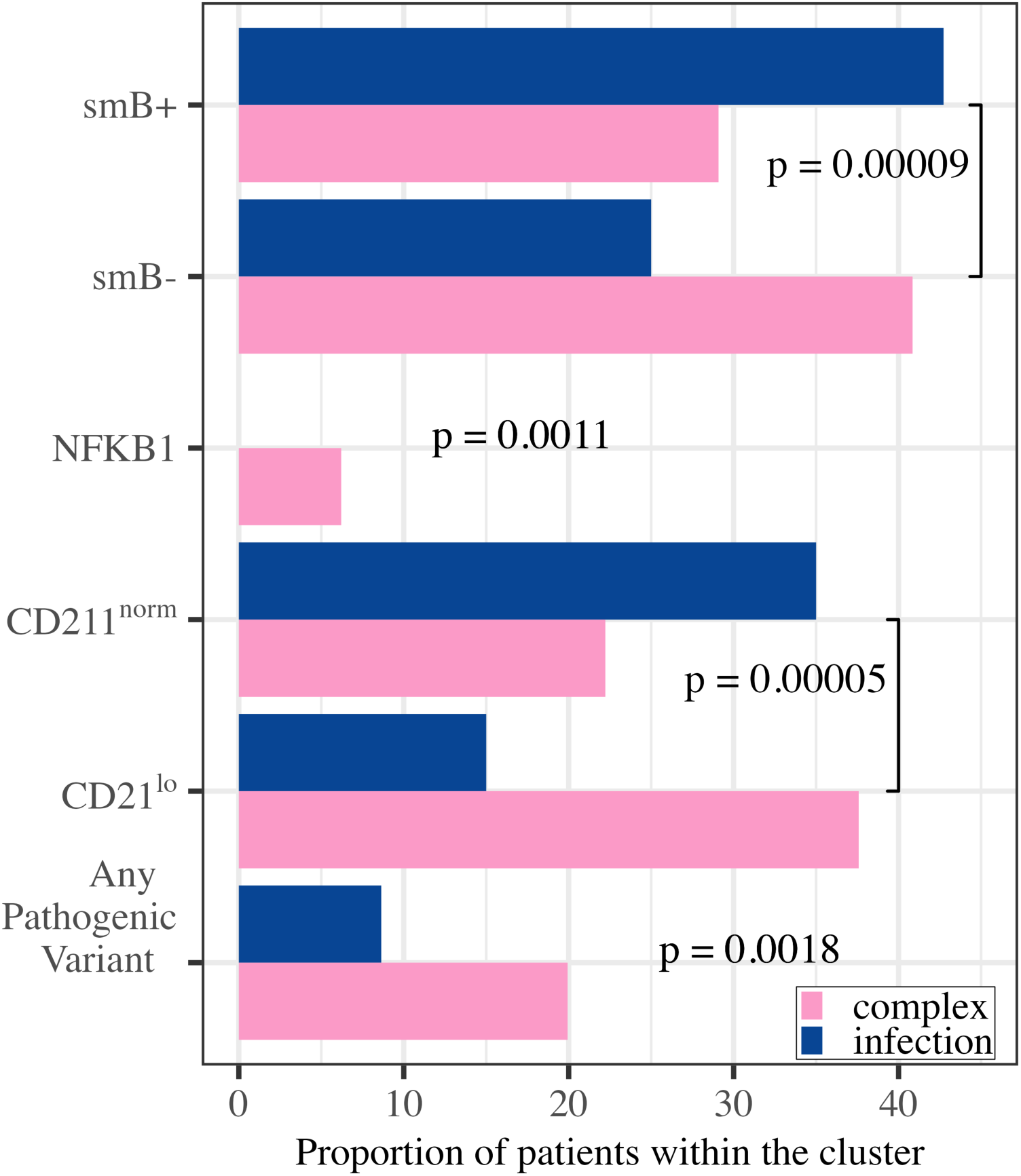
Complex patients more likely to have smB<2%, CD21^lo^>10%, and pathogenic variants including NFKB1 variants. Significant associations detected using Cochran-Mantel-Haenszel tests, stratified by contributing center to control for site-specific variation. *P* values were BH-adjusted within each test group.

### Phenotypic correlates of humoral and genetic variation in CVID

Patients with preserved IgA were significantly enriched for *Partial absence of specific antibody response to unconjugated pneumococcal vaccine* [HP:0410301] (BH adjusted *P* = 0.004687), while those with elevated IgM showed a strong association with *Pulmonary interstitial lymphocytic infiltration* [HP:0033582] (BH adjusted *P* = 4.5×10^-07^).

Among B cell parameters, very low levels of smB cells were significantly associated with splenomegaly (BH adjusted *P* = 0.0006). Expansion of the CD21^low^ subset was linked to autoimmunity (BH adjusted *P* = 0.017) and specifically to *Autoimmune thrombocytopenia* [HP:0001973] (BH adjusted *P* = 0.002241).

Regarding genetic correlates, canonical *TNFRSF13B (TACI)* variants were associated with *Recurrent candida infections* [HP:0005401] (BH adjusted *P* = 0.046). Variants in *NFKB1* were associated with *Autoimmune neutropenia* [HP:0001904] (BH adjusted *P* = 7.4×10^-6^). Pathogenic variants in other IUIS genes were associated with increased frequency of *Autoimmune hemolytic anemia* [HP:0001890] (BH adjusted *P* = 0.0091).

Table III summarises all statistically significant associations between immunoglobulin patterns, B cell subsets, genes, and specific HPO terms.

**Table III:**
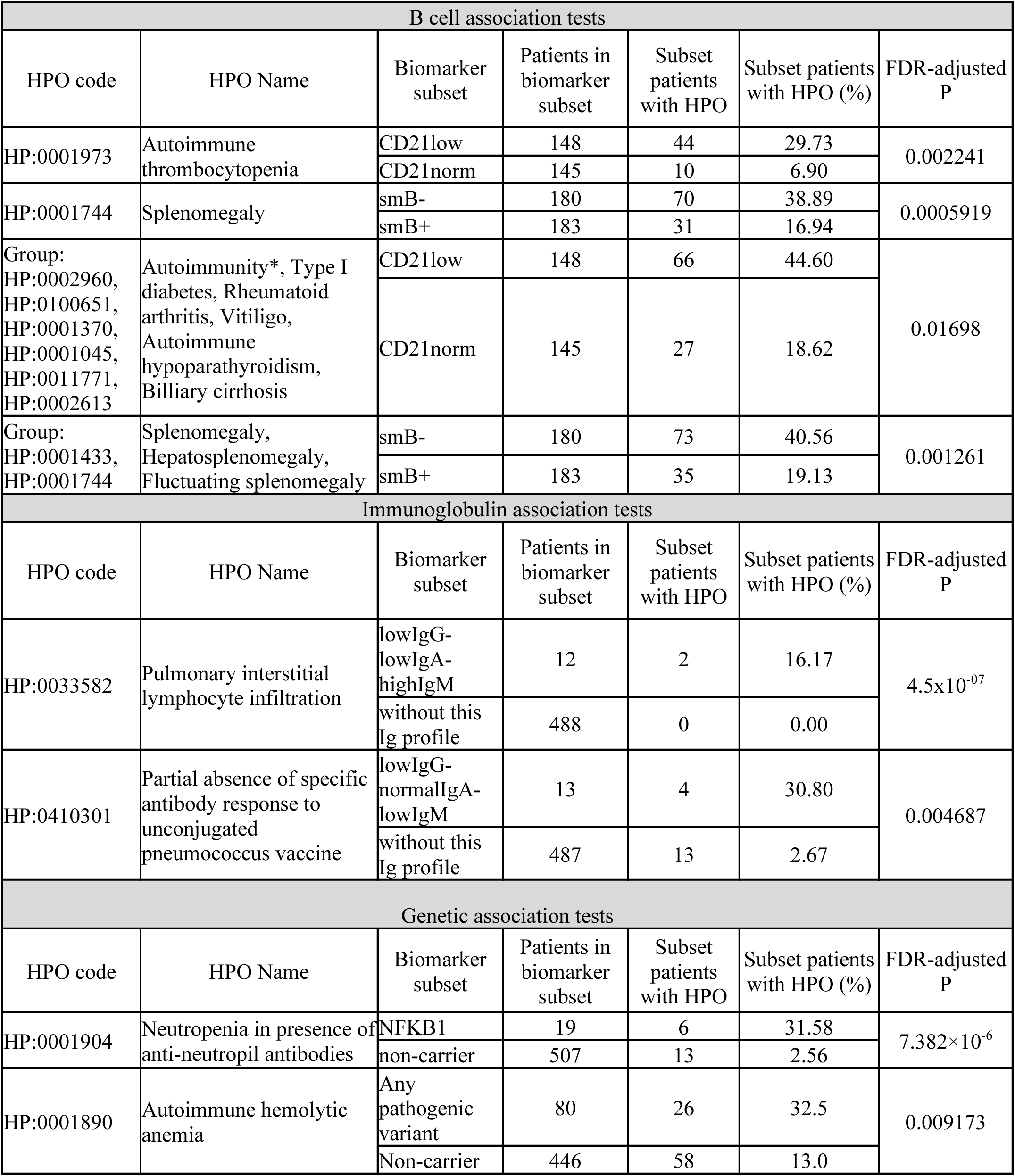
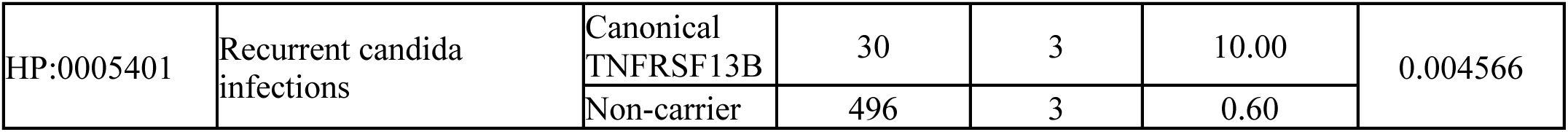
Rich clinical dataset allows granular investigation of associated patterns. Significant associations identified using Cochran-Mantel-Haenszel tests stratified by center. P values are BH-adjusted within each comparison group (B cell subtypes and HPO terms; Immunoglobulin profiles and HPO terms, Genetic diagnostic and HPO terms). For each biomarker, the patient count reflects only centers with at least one patient of the corresponding contingency table, ensuring the assumptions of the Cochran-Mantel-Haenszel test are met.

## Discussion

Deep clinical phenotyping is key to translational research and advancing genetic diagnosis. Our findings show that HPO is a feasible and effective approach for achieving this. It allows both individual-level comparisons – matching patient’s profile to known disease descriptions or other patients - and cohort-level analyses, to identify patterns, shared traits and distinct subgroups. Specifically, our findings support the application of HPO in CVID research.

Large, multicentre, well-characterised, HPO-coded cohorts have not yet been published for patients with PID, including CVID. Our study addresses this gap, providing what is, to our knowledge, the first large-scale application of HPO annotation to patients with PID. Our approach, combining a customised PCT with structured clinician training, substantially improved completeness, specificity, and consistency of HPO annotations and generated a dataset that is internally consistent and interoperable across platforms.

We expect that application of HPO in CVID will be beneficial. While large registries, such as ESID-R, remain essential for capturing information about human disease, heterogeneity in data depth and restricted quality control limit downstream analyses ^23^. Evidence from other disease cohorts demonstrates that deeply curated HPO annotation improves diagnostic yield ^27,28^. In systemic autoinflammatory diseases, structured phenotyping increased correct diagnoses from 66% to 86%, improving WGS prioritisation, and reducing candidate disease lists from 35 to 2^29^.

Our consistent phenotypic coding allowed categorisation of patients into clinically relevant disease groups – infection-only or complex CVID – when curating phenotype-defining term sets including both parent and descendent terms across relevant branches. This led to meaningful associations between B cell profiles, and pathogenic/likely pathogenic variants and disease complexity. Distinct associations were observed between B cell parameters and specific clinical phenotypes. Very low levels of switched memory B cells were significantly associated with splenomegaly, while expansion of the CD21^low^ B cell subset was linked to autoimmunity, particularly autoimmune thrombocytopenia. However, some of the associations previously reported by the well-known EUROClass study were not reproduced. Differences in cohort composition may in part explain this. Although the prevalence of autoimmune cytopenias and granulomatous disease was similar between cohorts (both approximately 20%), autoimmunity was more common (29.0% vs 20.3%), whereas splenomegaly and lymphadenopathy were less frequent in our cohort (27.7% vs 40.5% and 18.8% vs 26.2%, respectively) (Table E3 in the Online Repository). Immunophenotypically, we observed more smb+CD21^lo^ and fewer smb-Tr^hi^ patients (Figure 2). Differences in data ascertainment, added granularity of HPO phenotyping and stricter centre-stratified statistics with FDR correction likely explain the lack of replication of some EUROClass associations.

Associations with immunoglobulin levels were also identified. Patients with preserved IgA were enriched for impaired responses to unconjugated pneumococcal vaccine, suggesting that some of these individuals may align more closely with specific antibody deficiency (SPAD) rather than classical CVID. Elevated IgM was associated with pulmonary interstitial lymphocytic infiltration, in line with previous work linking increased IgM was linked with a higher risk for progressive interstitial lung disease ^30,31^ and autoimmunity. Autoimmune features were also previously linked to expansion of CD21low B cells ^32^.

CVID is an initial clinical label for patients, and participants in this study were compiled based on their CVID-like phenotype. Subsequent WGS identified monogenic disorders in a subset of patients. Given our sample size, we focused our analyses of HPO-gene variant associations on patients with *TACI* and *NFKB1* variants. Canonical *TNFRSF13B variants* were enriched in patients with *Recurrent candida infections* [HP:0005401], although this may be influenced by the small sample size. *NFKB1* variants, recognised as a major monogenic cause of CVID ^33^, were associated here with autoimmune neutropenia, and appear as a marker of complex disease, confirming reports that heterozygous *NFKB1* carriers show prominent immune dysregulation relative to other CVID cases ^34–38^.

Beyond demonstrating improved data quality, our findings underscore an often-overlooked challenge in ontology-based phenotyping: the HPO hierarchy can directly influence which patients are captured in computational analyses. Closely related clinical entities may be distributed across different branches of the ontology, only intersecting at the high-level node term *Phenotypic abnormality* [HP:0000118]. As a result, phenotypically similar patients may be overlooked unless queries are explicitly designed to traverse the full hierarchy, with targeted inclusion of clinically relevant terms. This process required deliberate review of the HPO structure to avoid missing patients annotated under non-overlapping branches and to accommodate clinically important features not yet represented in HPO, such as GLILD. These considerations highlight the relevance of our approach rather than relying solely on unsupervised hierarchical clustering, which does not consistently yield clinically interpretable groupings.

A limitation of our study arises from the patient datasets available for HPO annotation. Much of the clinically relevant information remains embedded in unstructured electronic health records, which is unlikely to be obtained with manual annotation. This hidden data represents an analytical gap that may be best addressed using technological advances in automated data extraction to enable deeper HPO annotation ^39,40^ and longitudinal data collection required for analysis and prediction of disease progression.

This will support research into the genetic architecture of CVID, including beyond single-gene causes, and help delineate mechanisms underlying disease subgroups.

Our study sets the scene for systematic, standardised phenotypic data collection to transform the study of CVID by shifting from a diagnosis-based framework to a truly phenotype-driven approach capable of revealing robust and unbiased associations. This will support research into the genetic architecture of CVID, including beyond single gene causes, and aid delineation of mechanisms that underly disease subgroups. Incorporating more detailed immunophenotyping, for example using high-parameter spectral flow cytometry that can characterise immune signatures at unprecedented granularity, is also likely to better enable biologically meaningful patient stratification that will direct and improve treatment of affected patients.

## Data Availability

All data produced in the present study are available upon reasonable request to the authors

## Abbreviations

CVID: Common variable immunodeficiency
HPO: Human phenotype ontology
IEI: Inborn error of immunity
PID: Primary immunodeficiency
WGS: Whole genome sequence
ESID: European Society for Immunodeficiencies
ERN-RITA: European Reference Network on Rare Immunodeficiency, Autoinflammatory and Autoimmune diseases
INTREPID: Integrative Translational Research in Primary immunodeficiency
PCT: Phenotype Capture Tool
NIHR: National Institute for Health and Care Research
MySQL: My Structured Query Language
SSL: Secure Sockets Layer
TLS: Transport Layer Security
PHP: Hypertext Preprocessor
IQR: Interquartile range
GLILD: Granulomatous lymphocytic interstitial lung disease
IUIS: International Union of Immunological Societies
*NFKB1*: Nuclear Factor Kapp B subunit 1
*TNFRSF13B*: Tumor Necrosis Factor Receptor Superfamily Member 13B
TACI: Transmembrane activator and CAML interactor
BH: Benjamini-Hochberg
FDR: False Discovery Rate
NHS: National Health Service
NRH: Nodular regenerative hyperplasia

## Methods

### Phenotype Capture tool (PCT) design

We developed the Phenotype Capture Tool (PCT) as part of the INTREPID project to replace previous paper-based data collection and facilitate the entry of detailed HPO terms for participants. The PCT was designed to be user-friendly and secure, and it allows data to be edited at any time to reflect the evolution of a patient’s clinical picture. The tool features an autocompletion HPO search box which displays a dynamic list of HPO terms and their synonyms as users type, enabling the selection of the most appropriate descriptors. It also includes fields for entering numerical laboratory values, as well as a free-text option for entering data not covered by existing HPO terms (Figure E1 in the Online Repository). The tool is written in PHP and JavaScript. All data are stored in a MySQL database on a virtual machine hosted on the University of Cambridge network. Data security is ensured by encrypted internet connection (SSL/TLS) and encrypted passwords. The database resides on a protected server that complies with general data protection regulation and institutional data-protection policies. Authentication is implemented through PHP sessions using encrypted passwords, and inactive users are automatically logged out.

In addition to the information entered by the clinicians and the full lists of HPO terms, the database stores information about the tool’s users, an event log and, optionally, a list of pre-approved patient IDs to guard against mistyping of IDs. This tracking ensures accountability, enhances data quality, and facilitates auditing, making the system more robust and reliable. Data can be downloaded as a csv file and visualised in R-generated automated plots, which are used for quality control purposes.

The PCT was initially populated with clinical and laboratory data collected from paper-based case report forms entered by clinicians at the time of patient recruitment to WGS. For this study, additional information was added to existing patient records by clinicians, following training as described below.

In addition to HPO terms, the following laboratory values were recorded for each participant: lymphocyte count, immunoglobulin levels, B cell subset counts including counts for naïve B cells, memory B cells, switched memory B cells (smB), CD21^low^ (CD21low) B cells, transitional B cells, and plasmablasts. All laboratory values were measured locally by the recruiting centres.

Records analysed in this study were contributed by 11 centres: Barts Health NHS Trust, Cambridge University Hospitals-Addenbrooke’s, Royal Papworth Hospital, Frimley Health NHS Foundation Trust, University Hospitals Birmingham NHS Foundation Trust, Leeds Teaching Hospitals NHS Trust, Queen Elizabeth University Hospital Glasgow, Royal Free Hospital, University Hospitals of North Midlands, Imperial College Healthcare, and Salford Royal NHS Foundation Trust.

### Clinician Training

Before accessing the PCT, clinicians at each recruiting centre received training on how to use the PCT, with emphasis on appropriate HPO term selection for patients with IEI. Training included practice using clinical case scenarios. In routine UK practice clinicians diagnose CVID according to the European Society for Immunodeficiencies (ESID) criteria, i.e., a marked decrease of IgG (at least 2 standard deviations below the mean for age) and a marked decrease in at least one of the isotypes IgM or IgA, the onset of clinical significant immunodeficiency at greater than 2 years of age, and the exclusion of defined causes of hypogammaglobulinemia ^23^.

Clinicians were reminded to apply ESID criteria when confirming CVID diagnosis.

### Patient categorisation

Patients were categorised based on the presence of HPO terms, after propagation of terms implied by the HPO’s *is-a* graph structure, according to a phenotype-defining set of HPO terms, including: Infection: *Unusual infection* [HP:0032101]; *Respiratory tract infection* [HP:0011947]; *Abscess* [HP:0025615; 2) Granulomas: *Granuloma* [HP:0032252; *Granulomatosis* [HP:0002955; *Pulmonary granulomatosis* [HP:0030250; 3) *Splenomegaly* [HP:0001744, 4) Lymphoproliferation: *Lymphadenopathy* [HP:0002716], *Lymphoid nodular hyperplasia* [HP:0011956], *Hematologic neoplasm* [HP:0004377]; 5) Autoimmunity: *Autoimmunity* [HP:0002960] plus *Type I diabetes* [HP:0100651], *Rheumatoid arthritis* [HP:0001370], *Primary biliary cirrhosis* [HP:0002613], *Autoimmune hypoparathyroidism* [HP:0011771] and *Vitiligo* [HP:0001045]; 6) Liver disease: *Nodular regenerative hyperplasia of liver* [HP:0011954]; 7) Bronchiectasis: *Bronchiectasis* [HP:0002110]. Presence or absence of granulomatous lymphocytic interstitial lung disease (GLILD) was recorded through a yes/no box in the PCT as GLILD is not an HPO term. Patients were classified into broad clinical groups – infection only or complex group – based on the presence or absence of non-infectious complications We classified patients according to their B cell subset counts relative to total B cells, following the EUROClass system ^11^: (1) smB-(≤ 2% smB) or smB+ (> 2%); (2) CD21^norm^ (CD21^low^ <10 %) or CD21^low^ (patients with expanded CD21^low^ subset ≥ 10%); (3) transitional B cell Tr^norm^ (<9%) or Tr^hi^ (≥ 9%).

Serum immunoglobulin levels categorised patients as: (1) low IgG (<7 g/L); (2) low (< 0.7 g/L) or normal (0.7-4 g/L) IgA; (3) low (<0.4g g/L) or normal (0.4 - 2.3 g/L) IgM. Panhypogammaglobulinaemia was defined as concomitantly low levels of all three isotypes (IgG, IgG, and IgM) according to the thresholds specified above.

We recorded pathogenic or likely pathogenic diagnostic variants in genes listed by the International Union of Immunological Societies (IUIS) ^26^. We separately analysed carriers of *NFKB1* variants, as well as canonical and non-canonical *TNFRSF13B (TACI)* variants, to assess stratified phenotypic patterns.

---

**Table E1:**
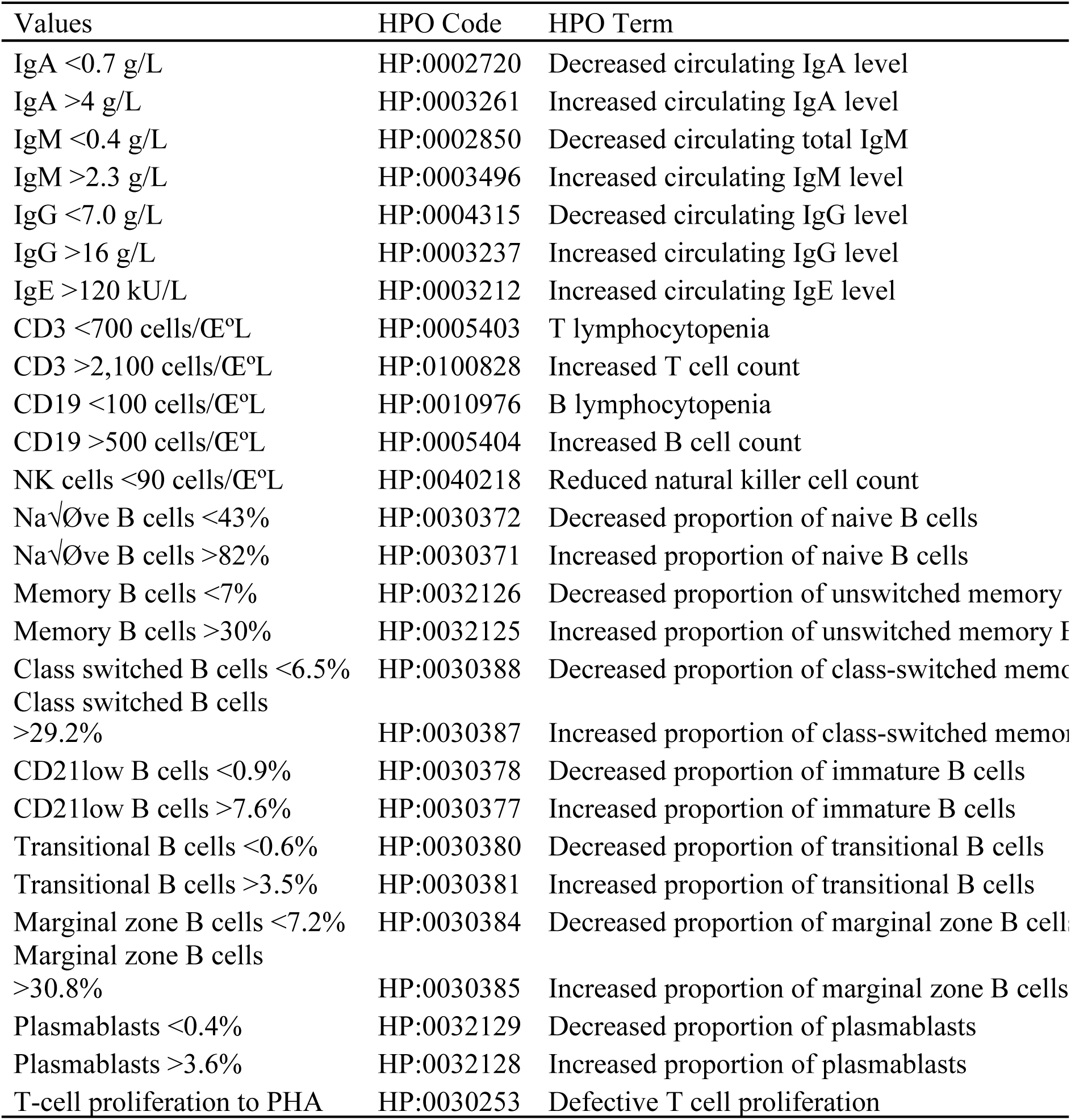
Mapping of laboratory values to HPO terms. Laboratory measurements were converted into corresponding HPO terms, Absolute CD4+ and CD8+ T cell counts are not shown, as no HPO terms currently exsit to describe absolute values; only proportional measurements. Terms describing absolute CD4+ and CD8+ counts are currently under submission to the HPO database.

**Table E2:**
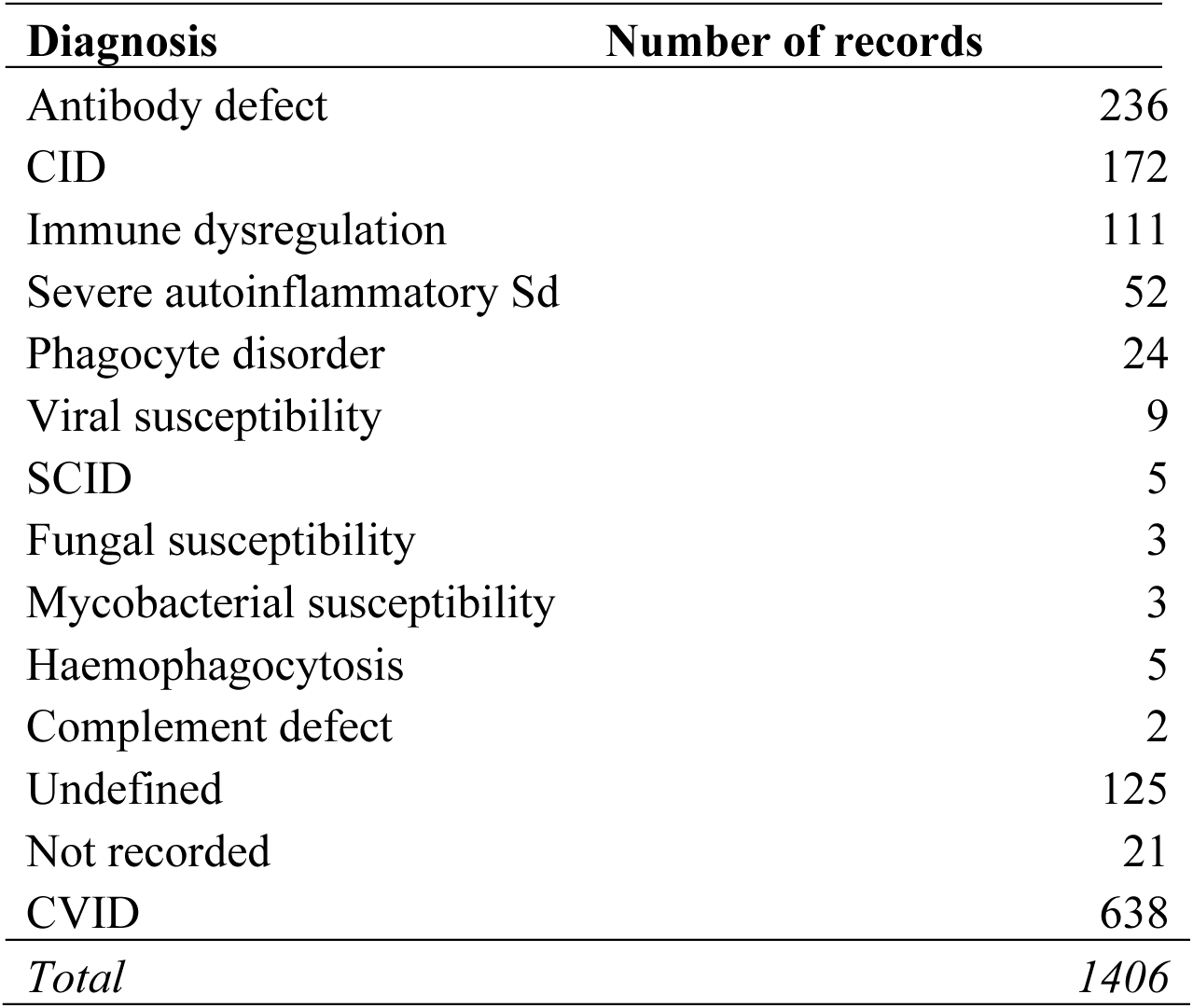
Distribution of Diagnosis Across the INTREPID CVID Cohort.

**Table E3:** Distribution of HPO Terms Across the INTREPID CVID Cohort.

| HPO code | Frequency | HPO term |
| --- | --- | --- |
| HP:0004315 | 516 | Decreased circulating IgG level |
| HP:0002720 | 470 | Decreased circulating IgA level |
| HP:0002850 | 420 | Decreased circulating total IgM |
| HP:0030388 | 342 | Decreased proportion of class-switched memory B cells |
| HP:0005363 | 224 | Humoral immunodeficiency |
| HP:0002110 | 217 | Bronchiectasis |
| HP:0010976 | 205 | B lymphocytopenia |
| HP:0002718 | 202 | Recurrent bacterial infections |
| HP:0040218 | 196 | Reduced natural killer cell count |
| HP:0032126 | 159 | Decreased proportion of unswitched memory B cells |
| HP:0002783 | 142 | Recurrent lower respiratory tract infections |
| HP:0001744 | 137 | Splenomegaly |
| HP:0005403 | 119 | T lymphocytopenia |
| HP:0030387 | 115 | Increased proportion of class-switched memory B cells |
| HP:0030384 | 107 | Decreased proportion of marginal zone B cells |
| HP:0032129 | 107 | Decreased proportion of plasmablasts |
| HP:0030371 | 94 | Increased proportion of naive B cells |
| HP:0005376 | 88 | Recurrent Haemophilus influenzae infections |
| HP:0001973 | 84 | Autoimmune thrombocytopenia |
| HP:0002788 | 83 | Recurrent upper respiratory tract infections |
| HP:0200117 | 80 | Recurrent upper and lower respiratory tract infections |
| HP:0005407 | 80 | Decreased proportion of CD4-positive helper T cells |
| HP:0030381 | 78 | Increased proportion of transitional B cells |
| HP:0002028 | 75 | Chronic diarrhea |
| HP:0410300 | 74 | Complete or near-complete absence of specific antibody response to unconjugated pneumococcus vaccine |
| HP:0030380 | 73 | Decreased proportion of transitional B cells |
| HP:0030377 | 73 | Increased proportion of immature B cells |
| HP:0006532 | 72 | Recurrent pneumonia |
| HP:0032125 | 70 | Increased proportion of unswitched memory B cells |
| HP:0002254 | 67 | Intermittent diarrhea |
| HP:0011109 | 63 | Chronic sinusitis |
| HP:0011108 | 63 | Recurrent sinusitis |
| HP:0005415 | 60 | Decreased proportion of CD8-positive T cells |
| HP:0005479 | 59 | Decreased circulating IgE |
| HP:0002910 | 57 | Elevated circulating hepatic transaminase concentration |
| HP:0045051 | 53 | Decreased DLCO |
| HP:0002099 | 52 | Asthma |
| HP:0003212 | 51 | Increased circulating IgE level |
| HP:0025179 | 50 | Ground-glass opacification |
| HP:0025392 | 43 | Nodular pattern on pulmonary HRCT |
| HP:0003155 | 40 | Elevated circulating alkaline phosphatase concentration |
| HP:0030385 | 38 | Increased proportion of marginal zone B cells |
| HP:0005404 | 38 | Increased B cell count |
| HP:0410295 | 38 | Complete or near-complete absence of specific antibody response to tetanus vaccine |
| HP:0030253 | 38 | Defective T cell proliferation |
| HP:0410018 | 38 | Recurrent ear infections |
| HP:0011954 | 37 | Nodular regenerative hyperplasia of liver |
| HP:0001890 | 37 | Autoimmune hemolytic anemia |
| HP:0002716 | 37 | Lymphadenopathy |
| HP:0002257 | 37 | Chronic rhinitis |
| HP:0001409 | 35 | Portal hypertension |
| HP:0001873 | 35 | Thrombocytopenia |
| HP:0000509 | 35 | Conjunctivitis |
| HP:0004429 | 34 | Recurrent viral infections |
| HP:0030372 | 32 | Decreased proportion of naive B cells |
| HP:0000010 | 32 | Recurrent urinary tract infections |
| HP:0004313 | 31 | Decreased circulating antibody level |
| HP:0000964 | 31 | Eczematoid dermatitis |
| HP:0033542 | 30 | Bronchial wall thickening |
| HP:0100721 | 27 | Mediastinal lymphadenopathy |
| HP:0011110 | 27 | Recurrent tonsillitis |
| HP:0033608 | 27 | Pulmonary nodule |
| HP:0045080 | 26 | Decreased proportion of CD3-positive T cells |
| HP:0001888 | 25 | Lymphopenia |
| HP:0001397 | 24 | Hepatic steatosis |
| HP:0000822 | 24 | Hypertension |
| HP:0001875 | 23 | Neutropenia |
| HP:0030378 | 23 | Decreased proportion of immature B cells |
| HP:0100828 | 23 | Increased T cell count |
| HP:0410303 | 22 | Complete or near-complete absence of specific antibody response to Haemophilus influenzae type b (Hib) vaccine |
| HP:0006510 | 22 | Chronic pulmonary obstruction |
| HP:0003460 | 21 | Decreased circulating total IgA |
| HP:0002040 | 21 | Esophageal varix |
| HP:0100502 | 21 | Decreased circulating vitamin B12 concentration |
| HP:0033374 | 21 | Decreased KCO |
| HP:0032342 | 21 | Reduced forced expiratory volume in one second |
| HP:0002206 | 20 | Pulmonary fibrosis |
| HP:0001904 | 19 | Neutropenia in presence of anti-neutrophil antibodies |
| HP:0005952 | 19 | obsolete Decreased pulmonary function |
| HP:0001903 | 18 | Anemia |
| HP:0002086 | 18 | Abnormality of the respiratory system |
| HP:0030250 | 18 | Pulmonary granulomatosis |
| HP:0002205 | 18 | Recurrent respiratory infections |
| HP:0005215 | 18 | Frequent Giardia lamblia infestation |
| HP:0030948 | 18 | Elevated gamma-glutamyltransferase level |
| HP:0001045 | 18 | Vitiligo |
| HP:0012476 | 18 | Decreased specific pneumococcal antibody level |
| HP:0012432 | 18 | Chronic fatigue |
| HP:0002758 | 18 | Osteoarthritis |
| HP:0002955 | 18 | Granulomatosis |
| HP:0002730 | 17 | Chronic noninfectious lymphadenopathy |
| HP:0410301 | 17 | Partial absence of specific antibody response to unconjugated pneumococcus vaccine |
| HP:0410299 | 17 | Decreased specific antibody response to polysaccharide vaccine |
| HP:0011473 | 17 | Villous atrophy |
| HP:0100827 | 17 | Lymphocytosis |
| HP:0008940 | 16 | Generalized lymphadenopathy |
| HP:0020099 | 16 | Severe norovirus infection |
| HP:0001891 | 16 | Iron deficiency anemia |
| HP:0002583 | 16 | Colitis |
| HP:0000821 | 16 | Hypothyroidism |
| HP:0001369 | 16 | Arthritis |
| HP:0003139 | 16 | Panhypogammaglobulinemia |
| HP:0002020 | 15 | Gastroesophageal reflux |
| HP:0002014 | 15 | Diarrhea |
| HP:0020106 | 15 | Severe giardiasis |
| HP:0012204 | 15 | Recurrent vulvovaginal candidiasis |
| HP:0410028 | 15 | Recurrent oral herpes |
| HP:0001081 | 15 | Cholelithiasis |
| HP:0002829 | 15 | Arthralgia |
| HP:0032341 | 15 | Reduced forced vital capacity |
| HP:0001438 | 14 | Abnormal abdomen morphology |
| HP:0011955 | 14 | Hepatic granulomatosis |
| HP:0011463 | 14 | Childhood onset |
| HP:0031123 | 14 | Recurrent gastroenteritis |
| HP:0001876 | 14 | Pancytopenia |
| HP:0005263 | 14 | Gastritis |
| HP:0033005 | 14 | Plantar warts |
| HP:0100750 | 14 | Atelectasis |
| HP:0002036 | 14 | Hiatus hernia |
| HP:0001882 | 14 | Leukopenia |
| HP:0033709 | 14 | Increased sputum production |
| HP:0031964 | 14 | Elevated circulating alanine aminotransferase concentration |
| HP:0012191 | 13 | B-cell lymphoma |
| HP:0005366 | 13 | Recurrent streptococcus pneumoniae infections |
| HP:0003496 | 13 | Increased circulating IgM level |
| HP:0002090 | 13 | Pneumonia |
| HP:0003765 | 13 | Psoriasiform dermatitis |
| HP:0011956 | 13 | Intestinal lymphoid nodular hyperplasia |
| HP:0000939 | 13 | Osteoporosis |
| HP:0100806 | 13 | Sepsis |
| HP:0012786 | 13 | Recurrent cystitis |
| HP:0025066 | 13 | Decreased mean corpuscular volume |
| HP:0003002 | 12 | Breast carcinoma |
| HP:0001287 | 12 | Meningitis |
| HP:0100280 | 12 | Crohn's disease |
| HP:0001433 | 12 | Hepatosplenomegaly |
| HP:0002608 | 12 | Celiac disease |
| HP:0005978 | 12 | Type II diabetes mellitus |
| HP:0002719 | 12 | Recurrent infections |
| HP:0025547 | 12 | Decreased mean corpuscular hemoglobin concentration |
| HP:0100512 | 12 | Decreased circulating vitamin D concentration |
| HP:0031965 | 12 | Increased RBC distribution width |
| HP:0031245 | 12 | Productive cough |
| HP:0005364 | 11 | obsolete Severe viral infections |
| HP:0001394 | 11 | Cirrhosis |
| HP:0006530 | 11 | Abnormal pulmonary interstitial morphology |
| HP:0200043 | 11 | Verrucae |
| HP:0001541 | 11 | Ascites |
| HP:0410305 | 11 | Partial absence of specific antibody response to Haemophilus influenzae type b (Hib) vaccine |
| HP:0032275 | 11 | Recurrent shingles |
| HP:0100658 | 11 | Cellulitis |
| HP:0006527 | 11 | Lymphocytic interstitial pneumonia |
| HP:0001250 | 10 | Seizure |
| HP:0100651 | 10 | Type I diabetes mellitus |
| HP:0410297 | 10 | Partial absence of specific antibody response to tetanus vaccine |
| HP:0000403 | 10 | Recurrent otitis media |
| HP:0000365 | 10 | Hearing impairment |
| HP:0002204 | 10 | Pulmonary embolism |
| HP:0002240 | 10 | Hepatomegaly |
| HP:0025390 | 10 | Reticular pattern on pulmonary HRCT |
| HP:0000872 | 9 | Hashimoto thyroiditis |
| HP:0032170 | 9 | Severe varicella zoster infection |
| HP:0002665 | 9 | Lymphoma |
| HP:0004798 | 8 | Recurrent infection of the gastrointestinal tract |
| HP:0012622 | 8 | Chronic kidney disease |
| HP:0003193 | 8 | Allergic rhinitis |
| HP:0020072 | 8 | Persistent EBV viremia |
| HP:0033004 | 8 | Palmar warts |
| HP:0025084 | 8 | Folliculitis |
| HP:0012475 | 8 | Decreased circulating level of specific antibody |
| HP:0002671 | 8 | Basal cell carcinoma |
| HP:0000246 | 8 | Sinusitis |
| HP:0011227 | 8 | Elevated circulating C-reactive protein concentration |
| HP:0025548 | 8 | Increased mean corpuscular hemoglobin concentration |
| HP:0003124 | 7 | Hypercholesterolemia |
| HP:0032128 | 7 | Increased proportion of plasmablasts |
| HP:0000155 | 7 | Oral ulcer |
| HP:0011949 | 7 | Acute infectious pneumonia |
| HP:0200123 | 7 | Chronic hepatitis |
| HP:0025289 | 7 | Cervical lymphadenopathy |
| HP:0030880 | 7 | Raynaud phenomenon |
| HP:0001061 | 7 | Acne |
| HP:0001370 | 7 | Rheumatoid arthritis |
| HP:0045042 | 7 | Decreased circulating complement C4 concentration |
| HP:0011370 | 7 | Recurrent cutaneous fungal infections |
| HP:0012384 | 7 | Rhinitis |
| HP:0000988 | 7 | Skin rash |
| HP:0100281 | 7 | Chronic colitis |
| HP:0001407 | 7 | Hepatic cysts |
| HP:0033628 | 7 | Bowel irritability |
| HP:0012378 | 7 | Fatigue |
| HP:0011123 | 6 | Inflammatory abnormality of the skin |
| HP:0002851 | 6 | Elevated proportion of CD4-negative, CD8-negative, alpha-beta regulatory T cells |
| HP:0033222 | 6 | Decreased CD4:CD8 ratio |
| HP:0033141 | 6 | Severe SARS-CoV-2 infection |
| HP:0003581 | 6 | Adult onset |
| HP:0032252 | 6 | Granuloma |
| HP:0002097 | 6 | Emphysema |
| HP:0002837 | 6 | Recurrent bronchitis |
| HP:0011919 | 6 | Pleural empyema |
| HP:0410302 | 6 | Decreased specific antibody response to protein-conjugated polysaccharide vaccine |
| HP:0001596 | 6 | Alopecia |
| HP:0011947 | 6 | Respiratory tract infection |
| HP:0100633 | 6 | Esophagitis |
| HP:0000498 | 6 | Blepharitis |
| HP:0002870 | 6 | Obstructive sleep apnea |
| HP:0005401 | 6 | Recurrent candida infections |
| HP:0005202 | 6 | Helicobacter pylori infection |
| HP:0100507 | 6 | Reduced blood folate concentration |
| HP:0003573 | 6 | Increased total bilirubin |
| HP:0030877 | 6 | Reduced FEV1/FVC ratio |
| HP:0030166 | 6 | Night sweats |
| HP:0032247 | 6 | Persistent CMV viremia |
| HP:0031956 | 6 | Elevated circulating aspartate aminotransferase concentration |
| HP:0000389 | 6 | Chronic otitis media |
| HP:0040089 | 6 | Abnormal natural killer cell count |
| HP:0003259 | 6 | Elevated circulating creatinine concentration |
| HP:0000787 | 5 | Nephrolithiasis |
| HP:0031393 | 5 | Abnormal proportion of CD8-positive T cells |
| HP:0002024 | 5 | Malabsorption |
| HP:0012203 | 5 | Onychomycosis |
| HP:0002037 | 5 | Inflammation of the large intestine |
| HP:0032216 | 5 | Lymphocytic infiltration of the colorectal mucosa |
| HP:0200118 | 5 | Malabsorption of Vitamin B12 |
| HP:0005365 | 5 | Severe B lymphocytopenia |
| HP:0002202 | 5 | Pleural effusion |
| HP:0002092 | 5 | Pulmonary arterial hypertension |
| HP:0002960 | 5 | Autoimmunity |
| HP:0410294 | 5 | Decreased specific antibody response to protein vaccine |
| HP:0100582 | 5 | Nasal polyposis |
| HP:0005266 | 5 | Intestinal polyp |
| HP:0006520 | 5 | Progressive pulmonary function impairment |
| HP:0002243 | 5 | Protein-losing enteropathy |
| HP:0006538 | 5 | Recurrent bronchopulmonary infections |
| HP:0002592 | 5 | Gastric ulcer |
| HP:0001738 | 5 | Exocrine pancreatic insufficiency |
| HP:0001395 | 5 | Hepatic fibrosis |
| HP:0001880 | 5 | Eosinophilia |
| HP:0004387 | 5 | Enterocolitis |
| HP:0032168 | 5 | Clostridium difficile colitis |
| HP:0032140 | 5 | Decreased specific antibody response to vaccination |
| HP:0033551 | 5 | Non-necrotizing pulmonary granulomatosis |
| HP:0032261 | 5 | Nontuberculous mycobacterial pulmonary infection |
| HP:0025435 | 5 | Increased circulating lactate dehydrogenase concentration |
| HP:0005110 | 5 | Atrial fibrillation |
| HP:0000716 | 5 | Depression |
| HP:0020059 | 5 | Increased red blood cell count |
| HP:0011107 | 4 | Recurrent aphthous stomatitis |
| HP:0030169 | 4 | Gastric varix |
| HP:0032167 | 4 | Clostridium difficile enteritis |
| HP:0032203 | 4 | Lymphoid nodular hyperplasia |
| HP:0031452 | 4 | Lichenoid skin lesion |
| HP:0005406 | 4 | Recurrent bacterial skin infections |
| HP:0011462 | 4 | Young adult onset |
| HP:0002582 | 4 | Atrophic gastritis |
| HP:0410133 | 4 | Chronic idiopathic urticaria |
| HP:0012125 | 4 | Prostate cancer |
| HP:0006775 | 4 | Multiple myeloma |
| HP:0033117 | 4 | Duodenitis |
| HP:0100665 | 4 | Angioedema |
| HP:0030374 | 4 | Decreased proportion of memory B cells |
| HP:0033657 | 4 | Linear atelectasis |
| HP:0001935 | 4 | Microcytic anemia |
| HP:0011897 | 4 | Neutrophilia |
| HP:0005231 | 4 | Chronic gastritis |
| HP:0006528 | 4 | Chronic lung disease |
| HP:0100575 | 4 | Neoplasm of the gallbladder |
| HP:0012701 | 4 | Bowel urgency |
| HP:0003095 | 4 | Septic arthritis |
| HP:0002253 | 4 | Colonic diverticula |
| HP:0003040 | 4 | Arthropathy |
| HP:0200008 | 4 | Intestinal polyposis |
| HP:0033125 | 4 | Follicular lymphoma |
| HP:0031207 | 4 | Hepatic hemangioma |
| HP:0000819 | 4 | Diabetes mellitus |
| HP:0005353 | 4 | Recurrent herpes |
| HP:0032218 | 4 | Decreased proportion of CD4-positive T cells |
| HP:0012539 | 4 | Non-Hodgkin lymphoma |
| HP:0002841 | 4 | Recurrent fungal infections |
| HP:0005425 | 4 | Recurrent sinopulmonary infections |
| HP:0031246 | 4 | Nonproductive cough |
| HP:0040126 | 4 | Abnormal circulating vitamin B12 concentration |
| HP:0020172 | 4 | Adverse drug response |
| HP:0020060 | 4 | Decreased red blood cell count |
| HP:0003270 | 4 | Abdominal distention |
| HP:0000951 | 3 | Abnormality of the skin |
| HP:0012189 | 3 | Hodgkin lymphoma |
| HP:0001658 | 3 | Myocardial infarction |
| HP:0012625 | 3 | Stage 3 chronic kidney disease |
| HP:0002664 | 3 | Neoplasm |
| HP:0012115 | 3 | Hepatitis |
| HP:0025400 | 3 | Nodular-random pattern on pulmonary HRCT |
| HP:0032242 | 3 | Cervical intraepithelial neoplasia |
| HP:0002232 | 3 | Patchy alopecia |
| HP:0012418 | 3 | Hypoxemia |
| HP:0009789 | 3 | Perianal abscess |
| HP:0001878 | 3 | Hemolytic anemia |
| HP:0006515 | 3 | Interstitial pneumonitis |
| HP:0020096 | 3 | Recurrent streptococcal infections |
| HP:0025389 | 3 | Pulmonary interstitial high-resolution computed tomography abnormality |
| HP:0100776 | 3 | Recurrent pharyngitis |
| HP:0012220 | 3 | Non-caseating epithelioid cell granulomatosis |
| HP:0012219 | 3 | Erythema nodosum |
| HP:0030991 | 3 | Sclerosing cholangitis |
| HP:0032219 | 3 | Increased proportion of CD4-positive T cells |
| HP:0030757 | 3 | Tooth abscess |
| HP:0002242 | 3 | Abnormal intestine morphology |
| HP:0000976 | 3 | obsolete Eczematoid dermatitis |
| HP:0003493 | 3 | Antinuclear antibody positivity |
| HP:0001581 | 3 | Recurrent skin infections |
| HP:0031949 | 3 | Recurrent bacterial upper respiratory tract infections |
| HP:0031292 | 3 | Cutaneous abscess |
| HP:0033636 | 3 | Combined pre- and post-capillary pulmonary hypertension |
| HP:0000554 | 3 | Uveitis |
| HP:0001698 | 3 | Pericardial effusion |
| HP:0100273 | 3 | Neoplasm of the colon |
| HP:0033351 | 3 | Candida esophagitis |
| HP:0100279 | 3 | Ulcerative colitis |
| HP:0032025 | 3 | Reduced circulating alpha-1-antitrypsin concentration |
| HP:0002754 | 3 | Osteomyelitis |
| HP:0001644 | 3 | Dilated cardiomyopathy |
| HP:0033280 | 3 | Paratracheal lymphadenopathy |
| HP:0002321 | 3 | Vertigo |
| HP:0000518 | 3 | Cataract |
| HP:0000360 | 3 | Tinnitus |
| HP:0010741 | 3 | Pedal edema |
| HP:0001328 | 3 | Specific learning disability |
| HP:0025127 | 3 | Actinic keratosis |
| HP:0000771 | 3 | Gynecomastia |
| HP:0010975 | 3 | Abnormal B cell count |
| HP:0002848 | 3 | Decreased specific anti-polysaccharide antibody level |
| HP:0001507 | 3 | Growth abnormality |
| HP:0032966 | 3 | Centrilobular emphysema |
| HP:0005518 | 3 | Increased mean corpuscular volume |
| HP:0031691 | 3 | Severe viral infection |
| HP:0002633 | 3 | Vasculitis |
| HP:0032386 | 3 | Elevated circulating transferrin concentration |
| HP:0011734 | 3 | Central adrenal insufficiency |
| HP:0002155 | 3 | Hypertriglyceridemia |
| HP:0005987 | 3 | Multinodular goiter |
| HP:0033176 | 3 | Submandibular lymph node enlargement |
| HP:0001824 | 3 | Weight loss |
| HP:0001971 | 3 | Hypersplenism |
| HP:0002721 | 3 | Immunodeficiency |
| HP:0001297 | 2 | Stroke |
| HP:0030078 | 2 | Lung adenocarcinoma |
| HP:0020103 | 2 | Invasive pulmonary aspergillosis |
| HP:0002728 | 2 | Chronic mucocutaneous candidiasis |
| HP:0009830 | 2 | Peripheral neuropathy |
| HP:0025615 | 2 | Abscess |
| HP:0005059 | 2 | Arthralgia/arthritis |
| HP:0010628 | 2 | Facial palsy |
| HP:0005162 | 2 | Abnormal left ventricular function |
| HP:0003774 | 2 | Stage 5 chronic kidney disease |
| HP:0004743 | 2 | Chronic tubulointerstitial nephritis |
| HP:0032301 | 2 | Genital warts |
| HP:0100723 | 2 | Gastrointestinal stroma tumor |
| HP:0005550 | 2 | Chronic lymphatic leukemia |
| HP:0012410 | 2 | Pure red cell aplasia |
| HP:0030049 | 2 | Brain abscess |
| HP:0010784 | 2 | Uterine neoplasm |
| HP:0020102 | 2 | Pneumocystis jirovecii pneumonia |
| HP:0550005 | 2 | Bilateral basilar pulmonary fibrosis |
| HP:0001970 | 2 | Tubulointerstitial nephritis |
| HP:0003261 | 2 | Increased circulating IgA level |
| HP:0100021 | 2 | Cerebral palsy |
| HP:0001631 | 2 | Atrial septal defect |
| HP:0033582 | 2 | Pulmonary interstitial lymphocyte infiltration |
| HP:0003459 | 2 | Polyclonal elevation of IgM |
| HP:0000992 | 2 | Cutaneous photosensitivity |
| HP:0002107 | 2 | Pneumothorax |
| HP:0002861 | 2 | Melanoma |
| HP:0033541 | 2 | Irreversible airflow obstruction |
| HP:0200029 | 2 | Vasculitis in the skin |
| HP:0032969 | 2 | Traction bronchiectasis |
| HP:0032294 | 2 | Monoclonal elevation of IgG kappa chain |
| HP:0032177 | 2 | Parenchymal consolidation |
| HP:0011945 | 2 | Bronchiolitis obliterans organizing pneumonia |
| HP:0002102 | 2 | Pleuritis |
| HP:0032972 | 2 | Nodular-centrilobular without tree-in-bud pattern on pulmonary HRCT |
| HP:0012486 | 2 | Myelitis |
| HP:0033000 | 2 | Subglottic laryngitis |
| HP:0002613 | 2 | Biliary cirrhosis |
| HP:0011850 | 2 | Parotitis |
| HP:0001101 | 2 | Iritis |
| HP:0025175 | 2 | Honeycomb lung |
| HP:0000421 | 2 | Epistaxis |
| HP:0031694 | 2 | Severe adenovirus infection |
| HP:0041063 | 2 | Chronic decreased circulating IgG2 |
| HP:0032174 | 2 | Tree-in-bud pattern |
| HP:0100769 | 2 | Synovitis |
| HP:0030692 | 2 | Brain neoplasm |
| HP:0005584 | 2 | Renal cell carcinoma |
| HP:0002604 | 2 | Gastrointestinal telangiectasia |
| HP:0004469 | 2 | Chronic bronchitis |
| HP:0002574 | 2 | Episodic abdominal pain |
| HP:0031944 | 2 | Pleural thickening |
| HP:0033540 | 2 | Reversible airflow obstruction |
| HP:0012122 | 2 | Anterior uveitis |
| HP:0009098 | 2 | Chronic oral candidiasis |
| HP:0032154 | 2 | Aphthous ulcer |
| HP:0005195 | 2 | Polyarticular arthropathy |
| HP:0033120 | 2 | Nummular eczema |
| HP:0001901 | 2 | Polycythemia |
| HP:0003326 | 2 | Myalgia |
| HP:0010638 | 2 | Elevated alkaline phosphatase of hepatic origin |
| HP:0000501 | 2 | Glaucoma |
| HP:0005145 | 2 | Coronary artery stenosis |
| HP:0025177 | 2 | Peribronchovascular interstitial thickening |
| HP:0003072 | 2 | Hypercalcemia |
| HP:0002027 | 2 | Abdominal pain |
| HP:0002904 | 2 | Hyperbilirubinemia |
| HP:0002076 | 2 | Migraine |
| HP:0001025 | 2 | Urticaria |
| HP:0004385 | 2 | Protracted diarrhea |
| HP:0012032 | 2 | Lipoma |
| HP:0010619 | 2 | Fibroadenoma of the breast |
| HP:0012387 | 2 | Bronchitis |
| HP:0025180 | 2 | Centrilobular ground-glass opacification on pulmonary HRCT |
| HP:0001019 | 2 | Erythroderma |
| HP:0001954 | 2 | Recurrent fever |
| HP:0004783 | 2 | Duodenal polyposis |
| HP:0002301 | 2 | Hemiplegia |
| HP:0000119 | 2 | Abnormality of the genitourinary system |
| HP:0001894 | 2 | Thrombocytosis |
| HP:0100796 | 2 | Orchitis |
| HP:0100577 | 2 | Urinary bladder inflammation |
| HP:0000093 | 2 | Proteinuria |
| HP:0012330 | 2 | Pyelonephritis |
| HP:0032305 | 2 | Decreased circulating mannose-binding protein concentration |
| HP:0003073 | 2 | Hypoalbuminemia |
| HP:0000726 | 2 | Dementia |
| HP:0410378 | 2 | Decreased proportion of naive CD4 T cells |
| HP:0032157 | 2 | Recurrent genital herpes |
| HP:0002019 | 2 | Constipation |
| HP:0025526 | 2 | Psoriasiform lesion |
| HP:0030337 | 2 | Elevated CD4-positive, CD25-positive regulatory T cell count |
| HP:0012626 | 2 | Stage 4 chronic kidney disease |
| HP:0025318 | 2 | Ovarian carcinoma |
| HP:0025173 | 2 | Nodular septal thickening on pulmonary HRCT |
| HP:0004398 | 2 | Peptic ulcer |
| HP:0000131 | 2 | Uterine leiomyoma |
| HP:0000107 | 2 | Renal cyst |
| HP:0003281 | 2 | Increased circulating ferritin concentration |
| HP:0025388 | 2 | Thyroid nodule |
| HP:0002021 | 2 | Pyloric stenosis |
| HP:0012450 | 2 | Chronic constipation |
| HP:0012343 | 2 | Decreased circulating ferritin concentration |
| HP:0000132 | 2 | Menorrhagia |
| HP:0000488 | 2 | Retinopathy |
| HP:0032551 | 2 | Hemorrhoids |
| HP:0000023 | 2 | Inguinal hernia |
| HP:0032259 | 2 | Chronic tinea infection |
| HP:0008223 | 2 | Compensated hypothyroidism |
| HP:0011040 | 2 | Abnormal intrahepatic bile duct morphology |
| HP:0030186 | 2 | Kinetic tremor |
| HP:0012184 | 2 | Increased HDL cholesterol concentration |
| HP:0003233 | 2 | Decreased HDL cholesterol concentration |
| HP:0001082 | 2 | Cholecystitis |
| HP:0033185 | 2 | Triggered by EBV infection |
| HP:0040303 | 2 | Decreased circulating iron concentration |
| HP:0410324 | 2 | Dust mite allergy |
| HP:0011948 | 2 | Recurrent acute respiratory tract infection |
| HP:0025408 | 2 | Abnormal spleen morphology |
| HP:0008843 | 2 | Hip osteoarthritis |
| HP:0001537 | 2 | Umbilical hernia |
| HP:0002315 | 2 | Headache |
| HP:0002588 | 2 | Duodenal ulcer |
| HP:0031394 | 2 | Abnormal CD4:CD8 ratio |
| HP:0002088 | 2 | Abnormal lung morphology |
| HP:0012735 | 2 | Cough |
| HP:0005424 | 2 | Absent specific antibody response |
| HP:0032016 | 2 | Abnormal sputum |
| HP:0003003 | 1 | Colon cancer |
| HP:0006555 | 1 | Diffuse hepatic steatosis |
| HP:0100646 | 1 | Thyroiditis |
| HP:0031700 | 1 | Invasive parasitic infection |
| HP:0002244 | 1 | Abnormal small intestine morphology |
| HP:0100617 | 1 | Testicular seminoma |
| HP:0012596 | 1 | Moderate proteinuria |
| HP:0002729 | 1 | Follicular hyperplasia |
| HP:0001300 | 1 | Parkinsonism |
| HP:0007417 | 1 | Discoid lupus rash |
| HP:0410325 | 1 | obsolete House dust mite allergy |
| HP:0000608 | 1 | Macular degeneration |
| HP:0410293 | 1 | Absent isohemagglutinin level |
| HP:0002863 | 1 | Myelodysplasia |
| HP:0030989 | 1 | Lymphoid cholangitis |
| HP:0002545 | 1 | Patchy demyelination of subcortical white matter |
| HP:0100583 | 1 | Corneal perforation |
| HP:0005561 | 1 | Abnormal bone marrow cell morphology |
| HP:0100616 | 1 | Testicular teratoma |
| HP:0007041 | 1 | Chronic lymphocytic meningitis |
| HP:0040328 | 1 | Focal hyperintensity of cerebral white matter on MRI |
| HP:0001272 | 1 | Cerebellar atrophy |
| HP:0100614 | 1 | Myositis |
| HP:0045049 | 1 | Abnormal DLCO |
| HP:0000265 | 1 | Mastoiditis |
| HP:0002722 | 1 | Recurrent abscess formation |
| HP:0003502 | 1 | Mild short stature |
| HP:0040275 | 1 | Adenocarcinoma of the large intestine |
| HP:0032577 | 1 | Clonal T cell receptor rearrangement |
| HP:0012192 | 1 | Cutaneous T-cell lymphoma |
| HP:0002586 | 1 | Peritonitis |
| HP:0001818 | 1 | Paronychia |
| HP:0003593 | 1 | Infantile onset |
| HP:0033327 | 1 | Nail psoriasis |
| HP:0040334 | 1 | Purulent rhinitis |
| HP:0011355 | 1 | Localized skin lesion |
| HP:0002094 | 1 | Dyspnea |
| HP:0003834 | 1 | Shoulder dislocation |
| HP:0012173 | 1 | Orthostatic tachycardia |
| HP:0008066 | 1 | Abnormal blistering of the skin |
| HP:0002308 | 1 | Chiari malformation |
| HP:0010562 | 1 | Keloids |
| HP:0033207 | 1 | Increased proportion autoreactive unresponsive CD21-/low B cells |
| HP:0001677 | 1 | Coronary artery atherosclerosis |
| HP:0002942 | 1 | Thoracic kyphosis |
| HP:0002756 | 1 | Pathologic fracture |
| HP:0032304 | 1 | Abnormal circulating mannose-binding protein concentration |
| HP:0007879 | 1 | Allergic conjunctivitis |
| HP:0008282 | 1 | Unconjugated hyperbilirubinemia |
| HP:0002113 | 1 | Pulmonary infiltrates |
| HP:0006719 | 1 | Benign gastrointestinal tract tumors |
| HP:0000419 | 1 | Abnormal nasal septum morphology |
| HP:0005420 | 1 | Recurrent gram-negative bacterial infections |
| HP:0020085 | 1 | Infection following live vaccination |
| HP:0025397 | 1 | Mosaic attenuation pattern on pulmonary HRCT |
| HP:0031284 | 1 | Flushing |
| HP:0000138 | 1 | Ovarian cyst |
| HP:0410017 | 1 | Otitis externa |
| HP:0033651 | 1 | Pulmonary mycetoma |
| HP:0005317 | 1 | Increased pulmonary vascular resistance |
| HP:0000938 | 1 | Osteopenia |
| HP:0001062 | 1 | Atypical nevus |
| HP:0010876 | 1 | Abnormal circulating protein concentration |
| HP:0002207 | 1 | Diffuse reticular or finely nodular infiltrations |
| HP:0012887 | 1 | Ovarian serous cystadenoma |
| HP:0007302 | 1 | Bipolar affective disorder |
| HP:0100586 | 1 | Sterile pyuria |
| HP:0020090 | 1 | Post-vaccination polio |
| HP:0031864 | 1 | Bacteremia |
| HP:0031498 | 1 | Mucinous gastric carcinoma |
| HP:0011892 | 1 | Decreased circulating vitamin K concentration |
| HP:0012190 | 1 | T-cell lymphoma |
| HP:0006536 | 1 | Airway obstruction |
| HP:0025044 | 1 | Lung abscess |
| HP:0033649 | 1 | Paraseptal emphysema |
| HP:0005264 | 1 | Abnormality of the gallbladder |
| HP:0001972 | 1 | Macrocytic anemia |
| HP:0025178 | 1 | Subpleural interstitial thickening |
| HP:0011274 | 1 | Recurrent mycobacterial infections |
| HP:0030058 | 1 | Sickled erythrocytes |
| HP:0032295 | 1 | Monoclonal elevation of IgG lambda chain |
| HP:0011967 | 1 | Decreased circulating copper concentration |
| HP:0002593 | 1 | Intestinal lymphangiectasia |
| HP:0002570 | 1 | Steatorrhea |
| HP:0012309 | 1 | Cutaneous amyloidosis |
| HP:0032134 | 1 | Chronic decreased circulating total IgG |
| HP:0100838 | 1 | Recurrent cutaneous abscess formation |
| HP:0200036 | 1 | Skin nodule |
| HP:0005517 | 1 | T-cell lymphoma/leukemia |
| HP:0004431 | 1 | Reduced circulating complement concentration |
| HP:0045038 | 1 | Gastric lymphoma |
| HP:0012490 | 1 | Panniculitis |
| HP:0100635 | 1 | Carotid paraganglioma |
| HP:0000105 | 1 | Enlarged kidney |
| HP:0012585 | 1 | Renal atrophy |
| HP:0033431 | 1 | Cytomegalovirus colitis |
| HP:0002326 | 1 | Transient ischemic attack |
| HP:0040289 | 1 | Cyclic neutropenia |
| HP:0033505 | 1 | Livedo reticularis |
| HP:0033663 | 1 | Air trapping |
| HP:0003390 | 1 | Sensory axonal neuropathy |
| HP:0000095 | 1 | Abnormal renal glomerulus morphology |
| HP:0005764 | 1 | Polyarticular arthritis |
| HP:0032645 | 1 | Renal interstitial mononuclear cell infiltration |
| HP:0000541 | 1 | Retinal detachment |
| HP:0100242 | 1 | Sarcoma |
| HP:0033584 | 1 | Nonspecific interstitial pneumonia |
| HP:0012089 | 1 | Arteritis |
| HP:0032661 | 1 | Generalized convulsive status epilepticus |
| HP:0005681 | 1 | Juvenile rheumatoid arthritis |
| HP:0033605 | 1 | Pustular rash |
| HP:0032643 | 1 | Renal interstitial non-necrotizing granulomas |
| HP:0001405 | 1 | Periportal fibrosis |
| HP:0030879 | 1 | Interlobular septal thickening |
| HP:0000704 | 1 | Periodontitis |
| HP:0000836 | 1 | Hyperthyroidism |
| HP:0009900 | 1 | Unilateral deafness |
| HP:0025085 | 1 | Bloody diarrhea |
| HP:0010769 | 1 | Pilonidal sinus |
| HP:0002017 | 1 | Nausea and vomiting |
| HP:0031842 | 1 | Lymphangiectasis |
| HP:0032124 | 1 | Abnormal proportion of unswitched memory B cells |
| HP:0002725 | 1 | Systemic lupus erythematosus |
| HP:0002091 | 1 | Restrictive ventilatory defect |
| HP:0000854 | 1 | Thyroid adenoma |
| HP:0006562 | 1 | Viral hepatitis |
| HP:0004800 | 1 | Duodenal diverticula |
| HP:0002625 | 1 | Deep venous thrombosis |
| HP:0031325 | 1 | Myocardial granulomatous infiltrates |
| HP:0012819 | 1 | Myocarditis |
| HP:0002607 | 1 | Bowel incontinence |
| HP:0001897 | 1 | Normocytic anemia |
| HP:0004386 | 1 | Gastrointestinal inflammation |
| HP:0031353 | 1 | Otitis media with effusion |
| HP:0002895 | 1 | Papillary thyroid carcinoma |
| HP:0100324 | 1 | Scleroderma |
| HP:0032408 | 1 | Breast mass |
| HP:0100534 | 1 | Episcleritis |
| HP:0000388 | 1 | Otitis media |
| HP:0200030 | 1 | Punctate vasculitis skin lesions |
| HP:0005236 | 1 | Chronic calcifying pancreatitis |
| HP:0032136 | 1 | Decreased circulating IgG1 level |
| HP:0100543 | 1 | Cognitive impairment |
| HP:0030871 | 1 | Facet joint arthrosis |
| HP:0012398 | 1 | Peripheral edema |
| HP:0410266 | 1 | Visceral hemangioma |
| HP:0033670 | 1 | Organizing pneumonia |
| HP:0005214 | 1 | Intestinal obstruction |
| HP:0012344 | 1 | Morphea |
| HP:0031801 | 1 | Vocal cord dysfunction |
| HP:0000870 | 1 | Increased circulating prolactin concentration |
| HP:0025232 | 1 | Bursitis |
| HP:0002925 | 1 | Elevated circulating thyroid-stimulating hormone concentration |
| HP:0000579 | 1 | Nasolacrimal duct obstruction |
| HP:0030127 | 1 | Endometriosis |
| HP:0000491 | 1 | Keratitis |
| HP:0008527 | 1 | Congenital sensorineural hearing impairment |
| HP:0410393 | 1 | Increased proportion of central memory CD4-positive, alpha-beta T cells |
| HP:0030351 | 1 | Urticarial plaque |
| HP:0025567 | 1 | Central serous chorioretinopathy |
| HP:0001289 | 1 | Confusion |
| HP:0000707 | 1 | Abnormality of the nervous system |
| HP:0002354 | 1 | Memory impairment |
| HP:0008165 | 1 | Decreased helper T cell proportion |
| HP:0000505 | 1 | Visual impairment |
| HP:0001382 | 1 | Joint hypermobility |
| HP:0000924 | 1 | Abnormality of the skeletal system |
| HP:0011946 | 1 | Bronchiolitis obliterans |
| HP:0003549 | 1 | Abnormality of connective tissue |
| HP:0002923 | 1 | Rheumatoid factor positive |
| HP:0001907 | 1 | Thromboembolism |
| HP:0001626 | 1 | Abnormality of the cardiovascular system |
| HP:0012443 | 1 | Abnormal brain morphology |
| HP:0008348 | 1 | Decreased circulating IgG2 level |
| HP:0020083 | 1 | Furuncle |
| HP:0002897 | 1 | Parathyroid adenoma |
| HP:0011114 | 1 | Defective production of NFkB1-dependent cytokines |
| HP:0008513 | 1 | Bilateral conductive hearing impairment |
| HP:0001999 | 1 | Abnormal facial shape |
| HP:0000598 | 1 | Abnormality of the ear |
| HP:0000044 | 1 | Hypogonadotropic hypogonadism |
| HP:0000975 | 1 | Hyperhidrosis |
| HP:0001260 | 1 | Dysarthria |
| HP:0033611 | 1 | Part-solid pulmonary nodule |
| HP:0025295 | 1 | Herpetiform |
| HP:0100580 | 1 | Barrett esophagus |
| HP:0006980 | 1 | Progressive leukoencephalopathy |
| HP:0000572 | 1 | Visual loss |
| HP:0004941 | 1 | Extrahepatic portal hypertension |
| HP:0001640 | 1 | Cardiomegaly |
| HP:0002858 | 1 | Meningioma |
| HP:0032557 | 1 | History of bone marrow transplant |
| HP:0004850 | 1 | Recurrent deep vein thrombosis |
| HP:0003011 | 1 | Abnormality of the musculature |
| HP:0001406 | 1 | Intrahepatic cholestasis |
| HP:0002902 | 1 | Hyponatremia |
| HP:0001955 | 1 | Unexplained fevers |
| HP:0002900 | 1 | Hypokalemia |
| HP:0002795 | 1 | Abnormal respiratory system physiology |
| HP:0012185 | 1 | Constrictive median neuropathy |
| HP:0011771 | 1 | Autoimmune hypoparathyroidism |
| HP:0006739 | 1 | Squamous cell carcinoma of the skin |
| HP:0008615 | 1 | Adult onset sensorineural hearing impairment |
| HP:0011950 | 1 | Bronchiolitis |
| HP:0004444 | 1 | Spherocytosis |
| HP:0005197 | 1 | Generalized morning stiffness |
| HP:0002183 | 1 | Phonophobia |
| HP:0000613 | 1 | Photophobia |
| HP:0000843 | 1 | Hyperparathyroidism |
| HP:0004755 | 1 | Supraventricular tachycardia |
| HP:0032152 | 1 | Keratosis pilaris |
| HP:0100726 | 1 | Kaposi's sarcoma |
| HP:0032641 | 1 | Renal interstitial granulomas |
| HP:0000820 | 1 | Abnormality of the thyroid gland |
| HP:0003141 | 1 | Increased LDL cholesterol concentration |
| HP:0001706 | 1 | Endocardial fibroelastosis |
| HP:0100822 | 1 | Rectocele |
| HP:0032215 | 1 | Disseminated cutaneous warts |
| HP:0001410 | 1 | Decreased liver function |
| HP:0002901 | 1 | Hypocalcemia |
| HP:0025346 | 1 | Increased circulating beta-2-microglobulin level |
| HP:0012117 | 1 | Hyperalbuminemia |
| HP:0031140 | 1 | Abnormal liver sonography |
| HP:0001737 | 1 | Pancreatic cysts |
| HP:0000083 | 1 | Renal insufficiency |
| HP:0002039 | 1 | Anorexia |
| HP:0000717 | 1 | Autism |
| HP:0002751 | 1 | Kyphoscoliosis |
| HP:0004322 | 1 | Short stature |
| HP:0100613 | 1 | Death in early adulthood |
| HP:0002293 | 1 | Alopecia of scalp |
| HP:0011952 | 1 | Acute aspiration pneumonia |
| HP:0001263 | 1 | Global developmental delay |
| HP:0000824 | 1 | Decreased response to growth hormone stimulation test |
| HP:0000252 | 1 | Microcephaly |
| HP:0001278 | 1 | Orthostatic hypotension |
| HP:0002153 | 1 | Hyperkalemia |
| HP:0003138 | 1 | Increased blood urea nitrogen |
| HP:0030828 | 1 | Wheezing |
| HP:0032326 | 1 | Methicillin-resistant Staphylococcus aureus infection |
| HP:0033350 | 1 | Elevated forced expiratory volume in one second |
| HP:0031384 | 1 | Reduced T cell CD40 expression |
| HP:0000407 | 1 | Sensorineural hearing impairment |
| HP:0006580 | 1 | Portal fibrosis |
| HP:0200120 | 1 | Chronic active hepatitis |
| HP:0001012 | 1 | Multiple lipomas |
| HP:0012431 | 1 | Episodic fatigue |
| HP:0001636 | 1 | Tetralogy of Fallot |
| HP:0002733 | 1 | Abnormal lymph node morphology |
| HP:0000729 | 1 | Autistic behavior |
| HP:0007018 | 1 | Attention deficit hyperactivity disorder |
| HP:0001513 | 1 | Obesity |
| HP:0032378 | 1 | Immediate-type hypersensitivity drug reaction |
| HP:0001028 | 1 | Hemangioma |
| HP:0025347 | 1 | Decreased circulating beta-2-microglobulin level |
| HP:0033590 | 1 | Inguinal abscess |
| HP:0033150 | 1 | Anorectal abscess |
| HP:0025196 | 1 | Increased total iron binding capacity |
| HP:0001197 | 1 | Abnormality of prenatal development or birth |
| HP:0033637 | 1 | Anti-endomysial antibody positivity |
| HP:0025546 | 1 | Abnormal mean corpuscular hemoglobin concentration |
| HP:0002875 | 1 | Exertional dyspnea |
| HP:0030874 | 1 | Oxygen desaturation on exertion |
| HP:0000739 | 1 | Anxiety |
| HP:0000712 | 1 | Emotional lability |
| HP:0100785 | 1 | Insomnia |
| HP:0001231 | 1 | Abnormal fingernail morphology |
| HP:0004299 | 1 | Hernia of the abdominal wall |
| HP:0012213 | 1 | Decreased glomerular filtration rate |
| HP:0041072 | 1 | Chronic decreased circulating IgG3 |
| HP:0001634 | 1 | Mitral valve prolapse |
| HP:0002650 | 1 | Scoliosis |
| HP:0020095 | 1 | Prolonged need of intravenous antibiotic therapy |
| HP:0410323 | 1 | Drug allergy |
| HP:0410149 | 1 | Drug-induced anaphylaxis |
| HP:0030151 | 1 | Cholangitis |
| HP:0008598 | 1 | Mild conductive hearing impairment |
| HP:0007099 | 1 | Chiari type I malformation |
| HP:0007017 | 1 | Progressive forgetfulness |
| HP:0005086 | 1 | Knee osteoarthritis |
| HP:0033687 | 1 | Short term memory impairment |
| HP:0040217 | 1 | Elevated hemoglobin A1c |
| HP:0012031 | 1 | Lipomatous tumor |
| HP:0020126 | 1 | Prostate mass |
| HP:0011593 | 1 | Left aortic arch with retroesophageal diverticulum of Kommerell |
| HP:0002015 | 1 | Dysphagia |
| HP:0001650 | 1 | Aortic valve stenosis |
| HP:0007141 | 1 | Sensorimotor neuropathy |
| HP:0031453 | 1 | Oral lichenoid lesion |
| HP:0041084 | 1 | Compression-fractured thoracic vertebra |
| HP:0005994 | 1 | Nodular goiter |
| HP:0001822 | 1 | Hallux valgus |
| HP:0410013 | 1 | Abnormality of the submandibular region |
| HP:0011915 | 1 | Cardiovascular calcification |
| HP:0002948 | 1 | Vertebral fusion |
| HP:0002573 | 1 | Hematochezia |
| HP:0033612 | 1 | Pure ground-glass pulmonary nodule |
| HP:0012312 | 1 | Monocytopenia |
| HP:0000100 | 1 | Nephrotic syndrome |
| HP:0012550 | 1 | Colonic varices |
| HP:0200063 | 1 | Colorectal polyposis |
| HP:0001735 | 1 | Acute pancreatitis |
| HP:0006824 | 1 | Cranial nerve paralysis |
| HP:0000651 | 1 | Diplopia |
| HP:0410321 | 1 | Animal dander allergy |
| HP:0000800 | 1 | Cystic renal dysplasia |
| HP:0001956 | 1 | Truncal obesity |
| HP:0003282 | 1 | Low alkaline phosphatase |
| HP:0004857 | 1 | Hyperchromic macrocytic anemia |
| HP:0012126 | 1 | Stomach cancer |
| HP:0025472 | 1 | Recurrent plantar mycosis |
| HP:0001387 | 1 | Joint stiffness |
| HP:0001717 | 1 | Coronary artery calcification |
| HP:0005679 | 1 | Dupuytren contracture |
| HP:0033343 | 1 | Mucoid diarrhea |
| HP:0025086 | 1 | Bloody mucoid diarrhea |
| HP:0000123 | 1 | Nephritis |
| HP:0011951 | 1 | Aspiration pneumonia |
| HP:0011499 | 1 | Mydriasis |
| HP:0032041 | 1 | Vocal cord polyp |
| HP:0000818 | 1 | Abnormality of the endocrine system |
| HP:0011116 | 1 | Abnormal circulating interferon concentration |
| HP:0002726 | 1 | Recurrent Staphylococcus aureus infections |
| HP:0008209 | 1 | Premature ovarian insufficiency |
| HP:0005268 | 1 | Miscarriage |
| HP:0004936 | 1 | Venous thrombosis |
| HP:0012317 | 1 | Sacroiliac arthritis |
| HP:0000230 | 1 | Gingivitis |
| HP:0033430 | 1 | Non-infectious meningitis |
| HP:0002018 | 1 | Nausea |
| HP:0012028 | 1 | Hepatocellular adenoma |
| HP:0001708 | 1 | Right ventricular failure |
| HP:0030243 | 1 | Hepatic vein thrombosis |
| HP:0100282 | 1 | Acute colitis |
| HP:0007717 | 1 | Chronic irritative conjunctivitis |
| HP:0010288 | 1 | Abnormality of the sublingual glands |
| HP:0030069 | 1 | Primary central nervous system lymphoma |
| HP:0001041 | 1 | Facial erythema |
| HP:0012636 | 1 | Retinal vein occlusion |
| HP:0031265 | 1 | Abnormal glomerular visceral epithelial cell morphology |
| HP:0004295 | 1 | Abnormal gastric mucosa morphology |
| HP:0008777 | 1 | Abnormal vocal cord morphology |
| HP:0410255 | 1 | Transient neutropenia |
| HP:0030955 | 1 | Addictive alcohol use |
| HP:0025223 | 1 | Triggered by smoking |
| HP:0012174 | 1 | Glioblastoma multiforme |
| HP:0001997 | 1 | Gout |
| HP:0012211 | 1 | Abnormal renal physiology |
| HP:0030838 | 1 | Hip pain |
| HP:0000364 | 1 | Hearing abnormality |
| HP:0004861 | 1 | Refractory macrocytic anemia |
| HP:0100526 | 1 | Neoplasm of the lung |
| HP:0020195 | 1 | IgG heavy chain paraproteinemia |
| HP:0000152 | 1 | Abnormality of head or neck |
| HP:0032522 | 1 | Ameliorated by immunosuppression |
| HP:0010474 | 1 | Bladder stones |
| HP:0001653 | 1 | Mitral regurgitation |
| HP:0008711 | 1 | Benign prostatic hyperplasia |
| HP:0008480 | 1 | Cervical spondylosis |
| HP:0100546 | 1 | Carotid artery stenosis |
| HP:0410281 | 1 | Dyspepsia |
| HP:0001681 | 1 | Angina pectoris |
| HP:0002761 | 1 | Generalized joint hypermobility |
| HP:0030386 | 1 | Abnormal proportion of class-switched memory B cells |
| HP:0033555 | 1 | Anti-Ro/SS-A antibody positivity |
| HP:0033016 | 1 | Chronic decreased circulating IgD |
| HP:0005523 | 1 | Lymphoproliferative disorder |
| HP:0001695 | 1 | Cardiac arrest |
| HP:0006771 | 1 | Duodenal adenocarcinoma |
| HP:0002246 | 1 | Abnormal duodenum morphology |
| HP:0100523 | 1 | Liver abscess |
| HP:0410243 | 1 | Abnormal circulating IgM level |
| HP:0000031 | 1 | Epididymitis |
| HP:0100561 | 1 | Spinal cord lesion |
| HP:0040270 | 1 | Impaired glucose tolerance |
| HP:0003763 | 1 | Bruxism |
| HP:0006740 | 1 | Transitional cell carcinoma of the bladder |
| HP:0004610 | 1 | Lumbar spinal canal stenosis |
| HP:0031952 | 1 | Neurogenic claudication |
| HP:0001097 | 1 | Keratoconjunctivitis sicca |
| HP:0000217 | 1 | Xerostomia |
| HP:0000077 | 1 | Abnormality of the kidney |
| HP:0012092 | 1 | Abnormality of exocrine pancreas physiology |
| HP:0011839 | 1 | Abnormal T cell count |
| HP:0031396 | 1 | Abnormal proportion of naive T cells |
| HP:0031379 | 1 | Abnormal T cell proliferation |
| HP:0032187 | 1 | Anal intraepithelial neoplasia |
| HP:0033221 | 1 | Increased CD4:CD8 ratio |
| HP:0001763 | 1 | Pes planus |
| HP:0001962 | 1 | Palpitations |
| HP:0100607 | 1 | Dysmenorrhea |
| HP:0003418 | 1 | Back pain |
| HP:0002970 | 1 | Genu varum |
| HP:0009725 | 1 | Bladder neoplasm |
| HP:0003489 | 1 | Acute episodes of neuropathic symptoms |
| HP:0410242 | 1 | Abnormal circulating IgG level |
| HP:0040268 | 1 | obsolete Recurrent infections of the middle ear |
| HP:0031352 | 1 | Chest tightness |
| HP:0005112 | 1 | Abdominal aortic aneurysm |
| HP:0030438 | 1 | Anal canal squamous cell carcinoma |
| HP:0030153 | 1 | Cholangiocarcinoma |
| HP:0032316 | 1 | Family history |
| HP:0100749 | 1 | Chest pain |
| HP:0001217 | 1 | Clubbing |
| HP:0001742 | 1 | Nasal congestion |
| HP:0002105 | 1 | Hemoptysis |
| HP:0031417 | 1 | Rhinorrhea |
| HP:0031261 | 1 | Bladder polyp |
| HP:0410376 | 1 | Increased proportion of naive CD8 T cells |
| HP:0031525 | 1 | Keratoacanthoma |

**Table E4:** Comparison between the INTREPID (n=526) and EUROClass (n=303) cohorts. Table summarizing cohort size, demographic composition, and frequencies of key clinical phenotypes in INTREPID compared with the EUROClass study.

|  | **INTREPID** | **EUROClass** |
| --- | --- | --- |
| Sex | 275 females, 243 males, 8 with no sex recorded | 169 females, 133 males |
| Year of birth (SD) | 1967 (17), 11 patients with no age recorded | 1957 (17) |
| Splenomegaly, % | 27.60% | 40.50% |
| Lymphadenopathy, % | 18.80% | 26.20% |
| Granulomatous disease, % | 10.10% | 11.60% |
| Autoimmunity, % | 29.10% | 20.30% |
| .... of which are Autoimmune cytopenias, % | 20% | 20.20% |
| NRH liver, % | 7% |  |
| Gastrointestinal inflammation, % | 9.90% |  |
| Hematological neoplasm, % | 7.60% |  |

**Figure E1:**
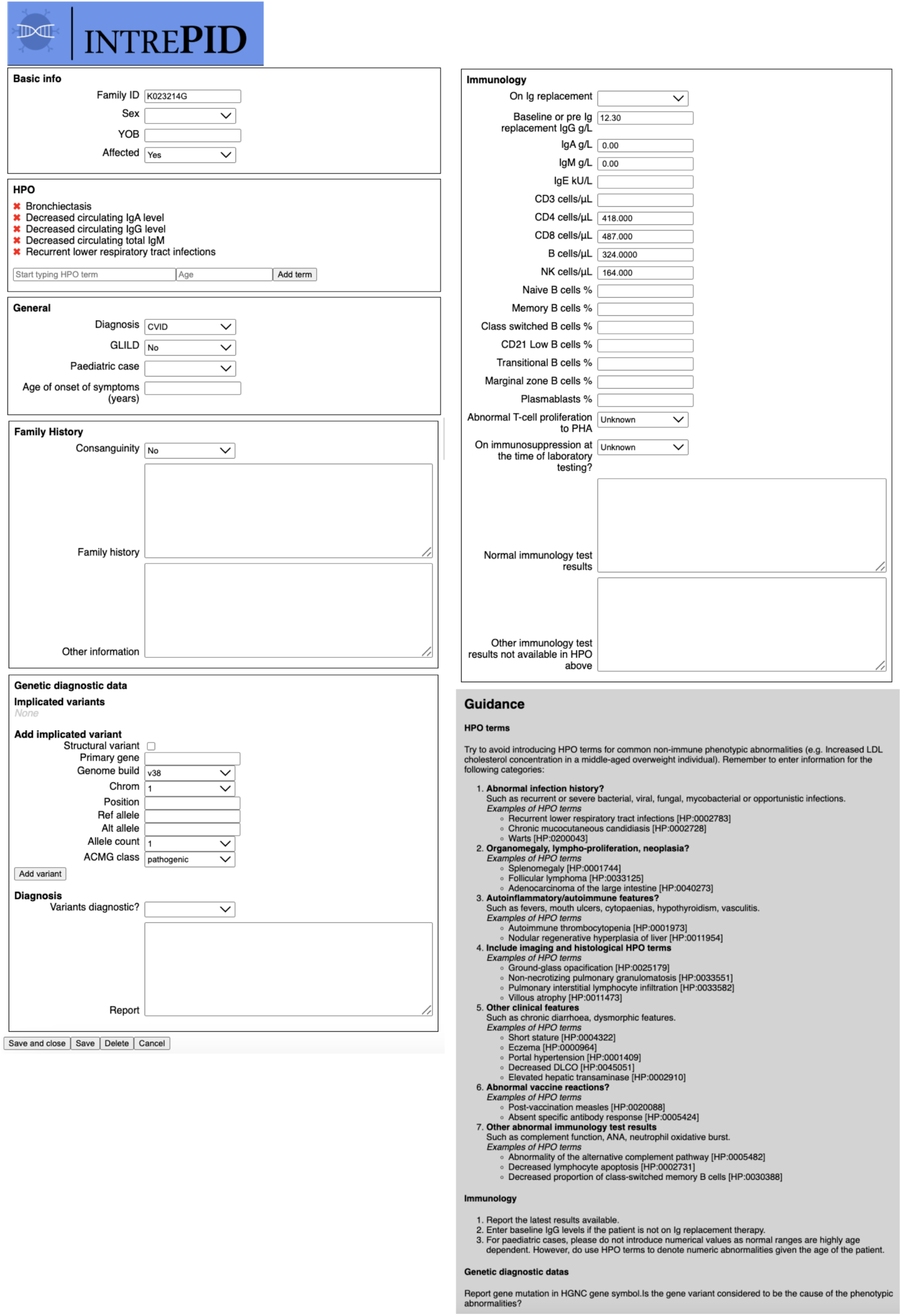
Phenotype Capture Tool (PCT). Interface used by clinicians to record structured phenotypic information using HPO.

**Figure E2:**
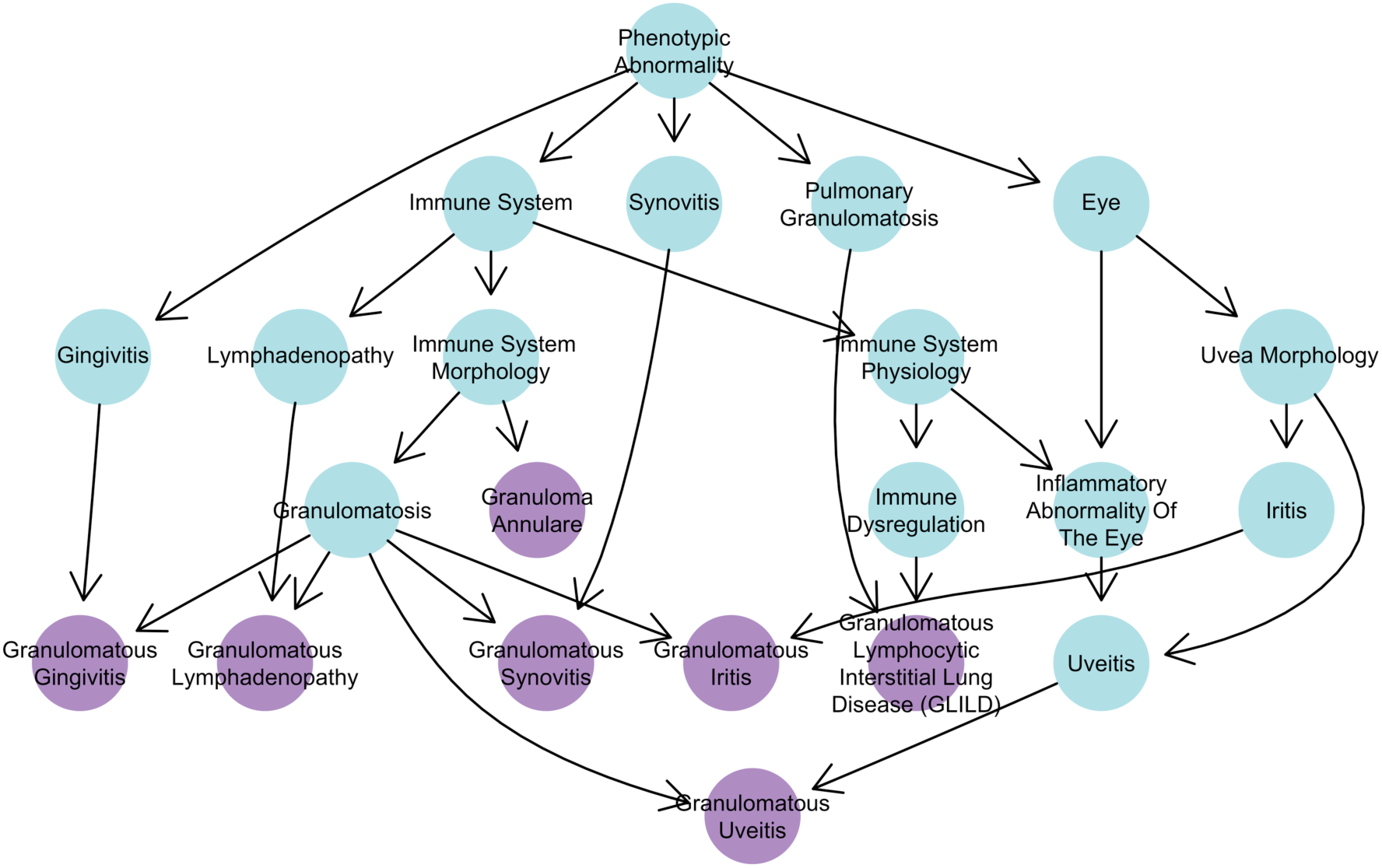
Directed acyclic graph illustrating proposed relationships and newly proposed granuloma-related HPO terms (purple)

